# Multi-organ post-acute sequelae of major respiratory and *Aedes*-borne arboviral diseases: a systematic review and meta-analysis

**DOI:** 10.64898/2026.05.15.26353287

**Authors:** Luis J Ponce, Baihui Xu, Esther Li Wen Choo, Jo Yi Chow, Rishi Rayapati, Bryan Zhi Ming Ling, Liang En Wee, Russ Li, David Chien Boon Lye, Eng Eong Ooi, Kelvin Bryan Tan, Jue Tao Lim

## Abstract

**Background:** Post-acute sequelae are well described following COVID-19 but may also occur after other respiratory infections and *Aedes*-borne infections. Evidence remains fragmented due to heterogeneity in study design, populations, and exposure, outcome, and follow-up definitions.

**Methods:** We synthesized and compared post-acute sequelae across influenza, RSV-ARI, dengue fever, chikungunya, Zika, and yellow fever. We searched five databases from inception to 25-08-2025 for articles quantifying risk, incidence, or rates of post-acute sequelae following these diseases. Eligible non-randomized observational studies assessed post-acute neurological, psychiatric, gastrointestinal, cardiovascular, respiratory, renal, musculoskeletal, autoimmune, or endocrine outcomes after confirmed infection. Risk of bias was assessed using ROBINS-E. Random-effects meta-analyses with restricted maximum likelihood estimation were conducted when ≥ 3 comparable effect estimates were available (PROSPERO #CRD420251124994).

**Findings:** 51 studies were included, predominantly from high-income regions. Most were retrospective cohorts using ICD-coded diagnoses; prospective studies used laboratory-confirmed infections. Data sources, comparator groups, exposure definitions, outcome ascertainment, and follow-up periods varied substantially. Meta-analyses were feasible for RSV, influenza, and dengue fever. All RSV-ARI studies were pediatric and assessed infections during infancy, which were associated with higher pooled odds of physician-diagnosed asthma (OR:2.93 [95%CI: 2.12–4.06]). Influenza studies used COVID-19-positive comparators; pooled estimates showed lower risk for neurological (HR:0.82 [0.76–0.89]) and composite outcomes (RR:0.88 [0.82–0.95]), with other organ systems non-significant. Dengue fever studies spanned all ages and showed increased risks of anxiety (HR:1.34 [1.01–1.78]), dementia (HR:1.61 [1.10–2.35]), autoimmune (RR:1.39 [1.17–1.67]), cardiovascular (HR:1.51 [1.27–1.80]), psychiatric (HR:1.17 [1.07–1.28]), and any sequelae (HR:1.19 [1.13–1.25]) versus those without prior infection.

**Interpretations:** Post-acute sequelae contribute to overall disease burden following RSV-ARI and dengue fever. The evidence remains limited by heterogeneity in study design, exposure and outcome definitions, comparator selection, and follow-up duration. Greater standardization in study design and reporting is needed to improve comparability and strengthen causal inference.

## Introduction

Post-acute sequelae of COVID-19 (PASC) has highlighted the potential disabling long-term consequences of viral infections. PASC is characterized by persistent symptoms such as fatigue, cognitive impairment and depressive symptoms lasting at least three months after the acute phase of illness,^1–3^ as well as increased risk of health conditions across multiple organ systems. Importantly, post-infectious chronic sequelae are not unique to COVID-19 and have been reported following recovery from a variety of acute viral infections, including other respiratory infections and *Aedes*-borne infections. Unlike other arboviral infections, *Aedes*-borne diseases are often more severe and widespread, and predominantly affect urban areas, leading to potentially large and consistent outbreaks in densely populated cities.^4,5^

Respiratory syncytial virus (RSV) and influenza virus infections pose significant public health concerns globally, especially among vulnerable populations.^6^ Influenza is a significant contributor in excess hospitalizations,^7^ while acute respiratory illness caused by RSV, included but not limited to bronchiolitis and pneumonia (hereon referred to as RSV-ARI), carries a high burden and morbidity globally.^8^ The acute burden of these illnesses is well established, with both being recognized as substantial contributors of global annual burden.^9,10^ Beyond the burden imposed during their infectious periods, there is growing evidence that longer-term complications may affect a significant proportion of patients as a result of influenza and RSV-ARI. Influenza, for example, is found to carry elevated risk of cardiovascular events post-infection, whereas RSV-ARI commonly causes increased risk of extra-pulmonary sequelae in adults.^11^ Despite growing recognition of post-acute sequelae (PAS) of viral infections, the current evidence base remains fragmented. For influenza and RSV-ARI, available studies have largely focused on selected outcomes (e.g., heart failure or asthma as complications), have been conducted in heterogeneous populations including only individuals exposed by RSV within their first one/two years of life, and have used inconsistent definitions and follow-up windows ranging from 4 weeks to several years post exposure, limiting comparability across settings.^12–15^

Elsewhere, *Aedes*-borne diseases’ substantial and growing global health burden are more apparent in tropical and subtropical regions where they disproportionately affect resource-limited populations.^16^ Vector-borne diseases may also lead to post-acute manifestations resulting in, for example, arthralgia, alopecia, or depression after Chikungunya infection, immunological and neuropsychiatric complications after dengue fever, or Guillain-Barre syndrome (GBS) after Zika fever.^17–20^ Yellow fever post-acute complications remain understudied, but it likely has some long-term complications following the pattern observed in other arboviral viruses. Although long-term manifestations after dengue fever have been documented, findings across studies vary considerably due to differences in study design, outcome ascertainment, and the timing of follow-up initiation and duration.^18,21,22^ Notably, evidence on PAS following chikungunya, Zika virus, and yellow fever remains sparse, and existing studies vary substantially in design, limiting comparability. Extracting the different study designs along with other study characteristics can inform which methods would be most practical to asses PAS for each viral infection.^23,24^ As a result of the lack of standardized evidence, the relative burden, spectrum, and temporal dynamics of post-acute sequelae across common respiratory and vector-borne viral infections remain poorly characterized.

To address these gaps, we conducted a systematic review and meta-analysis with the objective of synthesizing existing evidence on PAS following influenza, RSV-ARI, and dengue fever, chikungunya, Zika, and yellow fever). Using a standardized analytical framework, we evaluated a broad spectrum of post-acute outcomes spanning multiple organ systems, including cardiovascular, neurological, psychiatric, endocrine, autoimmune, and respiratory complications. We further assessed severe endpoints including all-cause hospitalization and mortality. In addition, selected symptom-based outcomes of clinical relevance were individually examined. Beyond data synthesis and interpretation, we also assessed every estimate’s risk of bias (RoB) across multiple domains with potential sources of bias, and pooled results that could be directly compared for each disease. These evaluations can also highlight methodological gaps and inconsistencies in study design, providing guidance for future research to adopt more standardized approaches that facilitate comparability and robust meta-analysis of PAS.

## Methods

### Systematic Review

We searched PubMed, Embase (via Ovid), CINAHL, Scopus, Web of Science, and Global Health from database inception to 25 August 2025, regardless of language. This review was conducted and reported in accordance with the Meta-analysis of Observational Studies in Epidemiology (MOOSE) guidelines,^25^ and the Preferred Reporting Items for Systematic Reviews and Meta-Analyses (PRISMA) was used to report the identification and screening procedure (Figure 1).^26^ Titles/abstracts and full texts were independently screened by two investigators (RR and BZML), with disagreements resolved by LJP. Forward citation was also performed. Full eligibility criteria details are described in Appendix C and Table S1. Search and data extraction details are described in Appendix C. This study was prospectively registered with PROSPERO (registration number: CRD420251124994).

**Figure 1:**
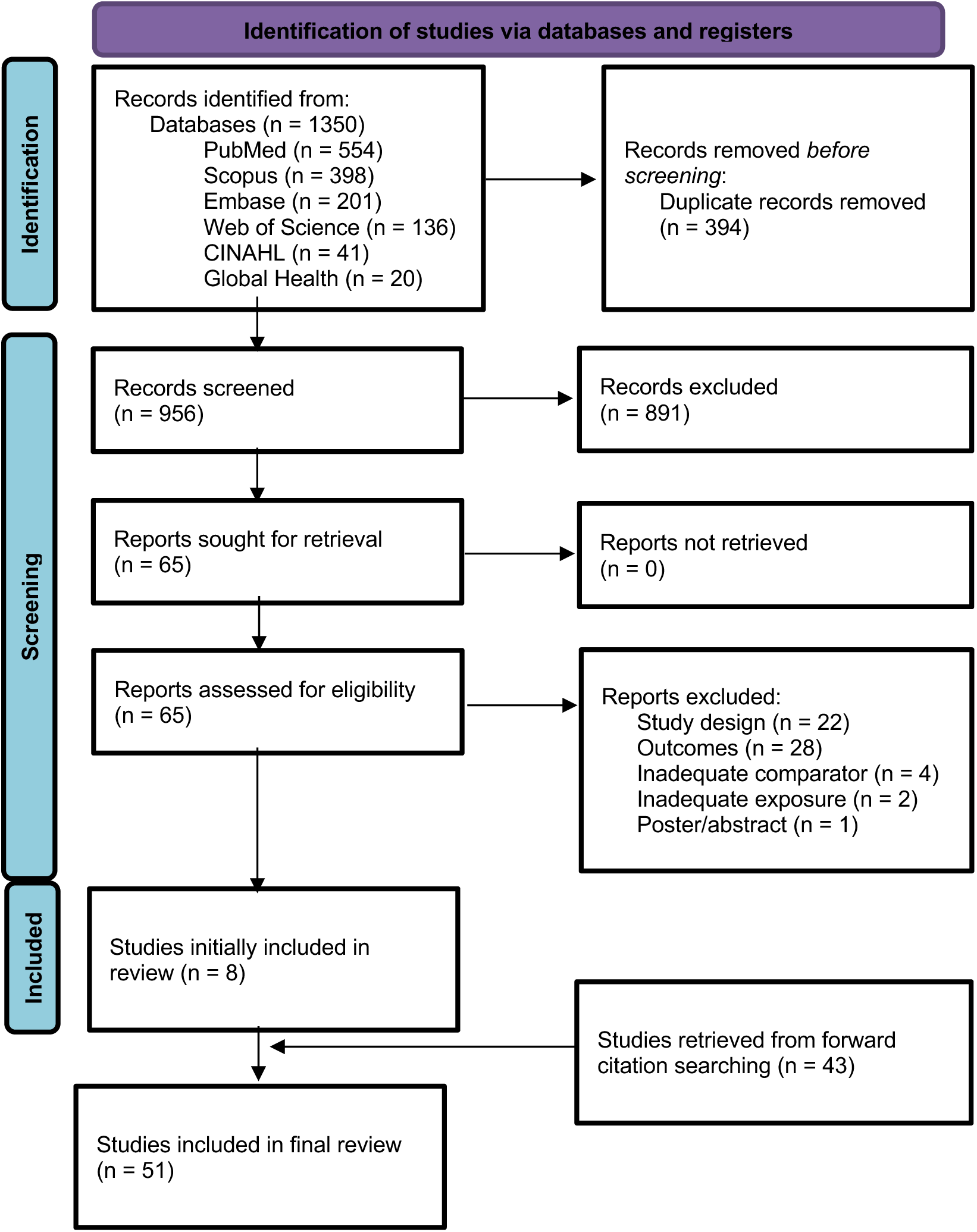
PRISMA flow diagram.

For quality assessment, we used the Risk Of Bias In Non-randomized Studies of Exposure (ROBINS-E) tool,^27^ treating each disease as the exposure and PAS, hospitalization, or death as outcomes. Studies were classified as low risk, some concerns, high risk, or very high risk of bias. RoB judgements were independently assigned by BX and LJP. Further RoB details and addressing of study appropriateness are described in Appendix C.

### Meta-analysis

For each of the six viruses included in this systematic review, we extracted data on incidence and effect estimates to conduct meta-analyses. Effect measures eligible for pooling included hazard ratios (HRs), incidence rate ratios (IRRs), and odds ratios (ORs). When the prevalence of the outcome in the control group or the baseline risk was known, we converted effect estimates and their confidence intervals to risk ratios (RRs) following the methodology proposed by Moser and McCann to convert HRs to RRs,^28^ and by Grant to convert ORs to RRs.^29^. When the prevalence of the outcome in the reference group is low (<10%), HRs and RRs are similar in magnitude. However, ORs tend to differ more substantially from their RR counterparts when outcomes are not rare, which warranted the conversions we performed.

Meta-analyses were conducted separately for each disease and outcome using comparable exposure–outcome models. Meta-analyses were performed only when at least three independent studies reported comparable estimates; otherwise, results were summarized descriptively. Random-effects models with restricted maximum likelihood estimation were used to account for between-study heterogeneity. Heterogeneity was assessed using τ² (the between-study variance in effect sizes), I² (the proportion of total variation attributable to true heterogeneity rather than sampling error), and H² (the relative excess of observed variability) statistics.^30^ As high heterogeneity is expected between studies, we also included the prediction interval for each meta-analysis.^31,32^ Potential publication bias was evaluated using funnel plots, which assess the association between study precision and effect size. Symmetry is expected in the absence of bias, whereas asymmetry may indicate publication bias or small-study effects.^33^ Sensitivity analyses were conducted excluding studies at high RoB. Further details regarding our meta-analyses are described in Appendix C.

## Results

51 studies met inclusion criteria: 8 identified from the primary search and 43 via forward citation searching. Studies were conducted across multiple global regions, predominantly in high-income settings, but distributed differently depending on the type of disease (respiratory versus *Aedes*-borne; Figure 2).

**Figure 2:**
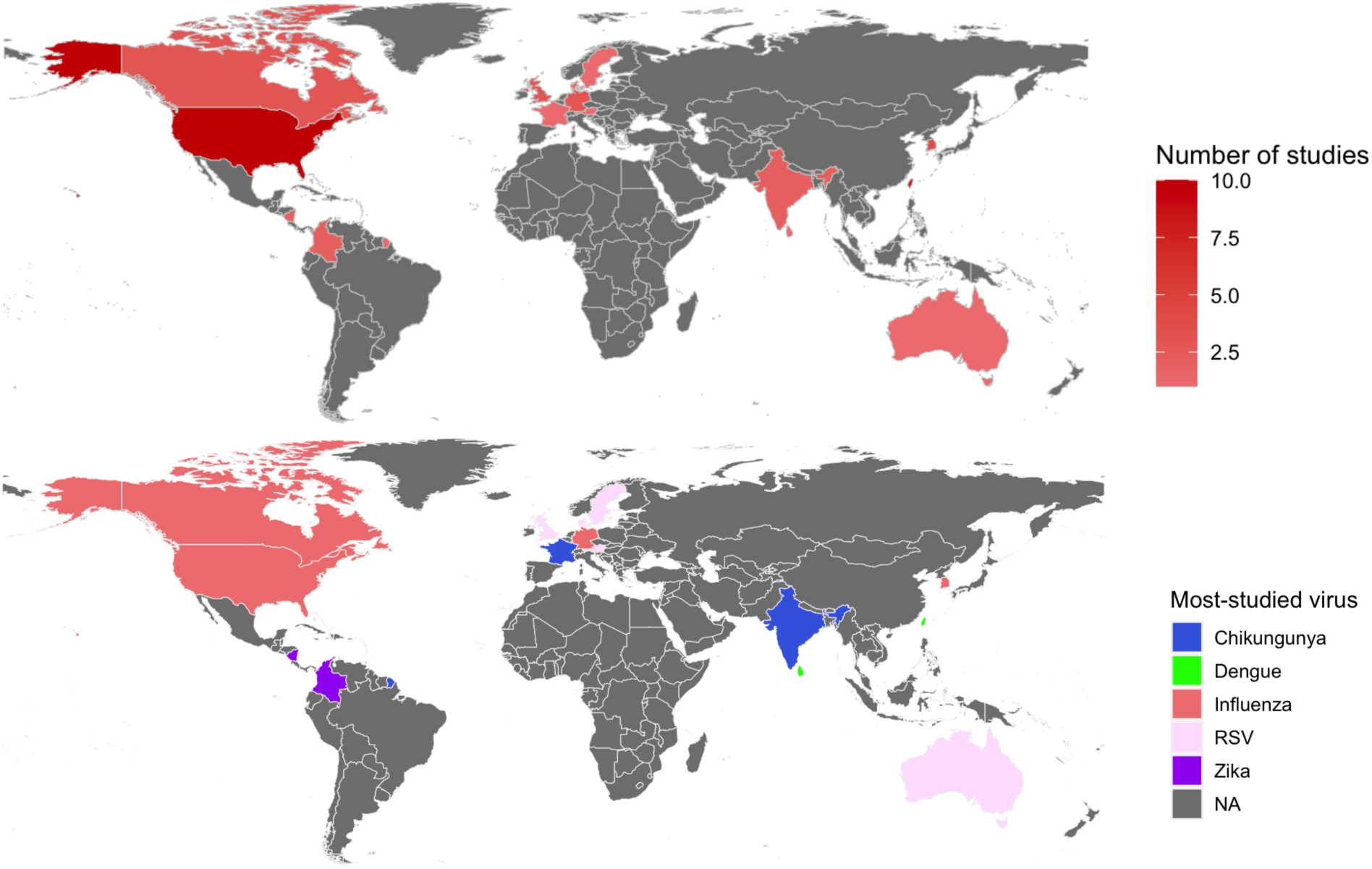
Maps of countries with studies included in this review and the disease with the higher number of studies per country.

Post-acute sequelae following dengue fever was more frequently studied than that of the other 5 diseases (16),^21,22,34–47^ followed by influenza (15),^44,53,57–59,65,66,68,70,78,80–82,82–84^ and symptomatic RSV infection (14).^11,15,63–74^ Zika fever and chikungunya disease were examined as exposures in four and three studies, respectively.^20,75–80^ No study with yellow fever as the exposure met the inclusion criteria for this analysis. Most studies were retrospective cohort designs (65%), followed by prospective cohort studies (27%), with few case-control (4%) or descriptive surveillance (4%) studies.

RSV-ARI studies primarily evaluated respiratory outcomes such as asthma, whereas dengue fever and influenza studies examined a broader range of multi-organ outcomes (Figure 3). Exposure definitions varied, with prospective studies typically using laboratory-confirmed diagnoses and retrospective studies relying on ICD-coded diagnoses. Chikungunya studies reported incidence of neurological outcomes (0.54%),^80^ rheumatoid arthritis (16.75%),^77^ and arthralgia (0.06%) among those with a history of disease.^78^ Zika studies reported incidences of any post-acute sequelae (0.39%, 3%, and 11%),^20,75,76^ all-cause mortality (0.03%),^20^ and neurological outcomes (3.3%).^79^ Individual and summary RoB assessments are shown in Table 1 and Supplementary Materials Figure S1 and S2.^81^

**Figure 3:**
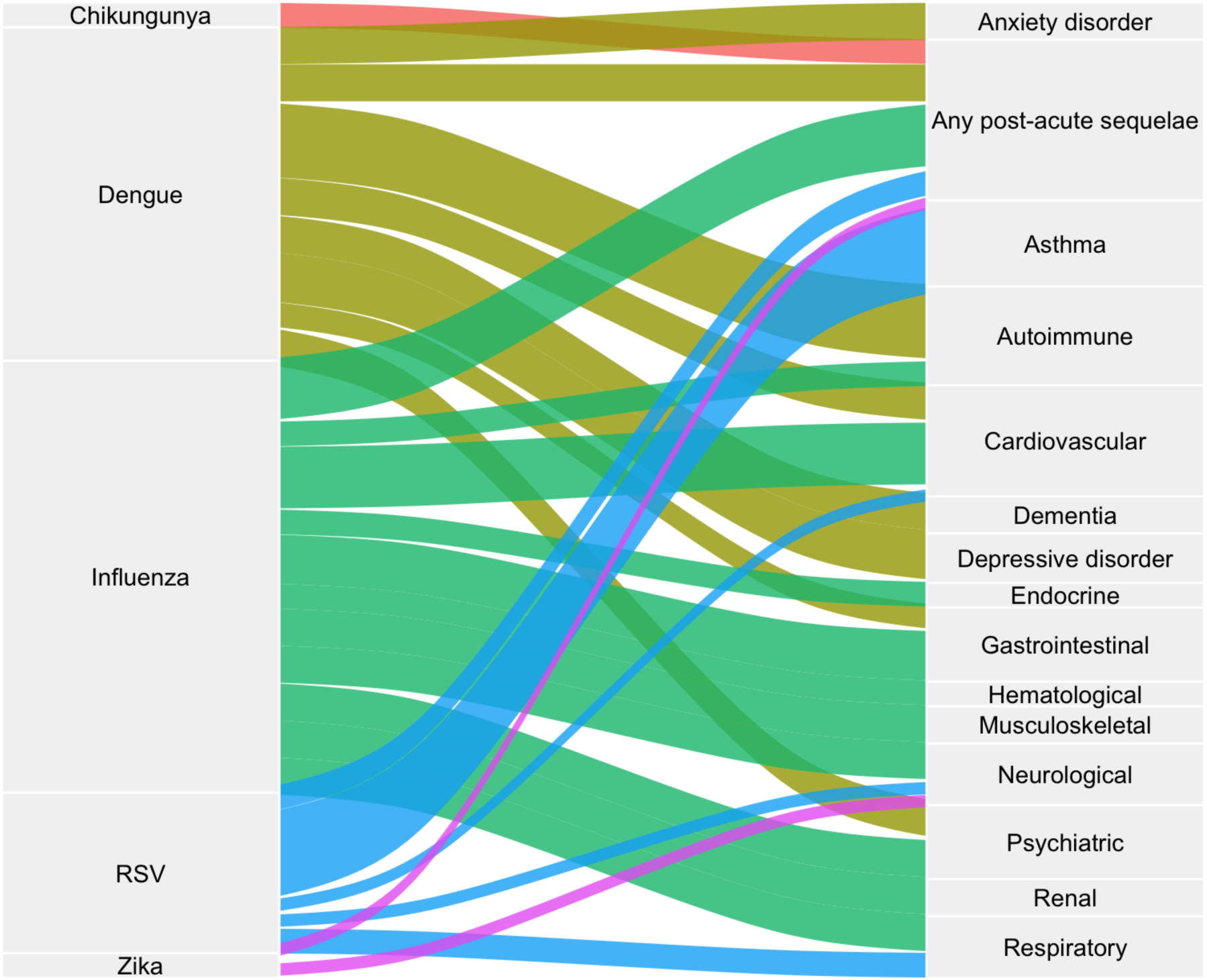
Mapping of viral exposures to post-acute outcomes by organ system.

**Table 1:** Demographics, study designs, study population characteristics, effect measures, and risks of bias for each study included in the review.

| Author(s) | Year | Country | Income Level | Disease | Study Design | Sample size: exposed | Sample size: control | Effect Measure | Risk of Bias |
| --- | --- | --- | --- | --- | --- | --- | --- | --- | --- |
| Liu et al | 2023 | Multinational | NA | Influenza | Retrospective cohort | 11266 | 7187 | Hazard ratio | High |
| Wee et al | 2025a | Singapore | High | Influenza | Retrospective cohort | 10454 | 70628 | Hazard ratio | Some concerns |
| Lim et al | 2025 | Singapore | High | Dengue fever | Retrospective cohort | 55870 | 3072309 | Hazard ratio | Some concerns |
| Wee et al | 2024 | Singapore | High | Dengue fever | Retrospective cohort | 11707 | 1248326 | Hazard ratio | Low |
| Wee et al | 2025b | Singapore | High | RSV | Retrospective cohort | 10193 | 76928 | Hazard ratio | Some concerns |
| Escobar et al | 2013 | USA | High | RSV | Retrospective cohort | 1263 | 71339 | Odds ratio | Some concerns |
| Iosifescu et al | 2022 | USA | High | Influenza | Retrospective cohort | 5772 | 18811 | Incidence | Did not pass appropriateness |
| Schaefer et al | 2025 | USA | High | Influenza | Retrospective cohort | 20844 | 121205 | Odds ratio | High |
| Li et al | 2018 | Taiwan | High | Dengue fever | Prospective cohort | 2506 | 112554 | Hazard ratio | Some concerns |
| Shih et al | 2024 | Taiwan | High | Dengue fever | Retrospective cohort | 45334 | 226670 | Hazard ratio | Some concerns |
| Shih et al | 2023 | Taiwan | High | Dengue fever | Retrospective cohort | 3814 | 255256 | Hazard ratio | Some concerns |
| Chien et al | 2023 | Taiwan | High | Dengue fever | Retrospective cohort | 37928 | 151712 | Hazard ratio | High |
| Wee et al | 2025 | Singapore | High | Dengue fever | Retrospective cohort | 6452 | 260749 | Hazard ratio | Low |
| Li et al | 2018 | Taiwan | High | Dengue fever | Retrospective cohort | 13787 | 13787 | Hazard ratio | Low |
| Xie et al | 2024 | USA | High | Influenza | Retrospective cohort | 10985 | 81280 | Hazard ratio | Some concerns |
| Bajema et al | 2025 | USA | High | Influenza | Retrospective cohort | 36189 | 289344 | Risk difference | Some concerns |
| Lewnard et al | 2025 | USA | High | Influenza | Retrospective cohort | 18790 | 74738 | Hazard ratio | Some concerns |
| Mejias et al | 2019 | USA | High | RSV | Retrospective cohort | 124439 | 124439 | Incidence rate ratio | High |
| Lu et al. | 2023 | USA | High | Influenza | Retrospective cohort | 2988 | 11214 | Odds ratio | Some concerns |
| Zhang et al. | 2023 | USA | High | Influenza | Retrospective cohort | 2547 | 7020 | Odds ratio | Very High |
| Fung et al. | 2023 | USA | High | Influenza | Retrospective cohort | 933877 | 2071959 | Odds ratio | Some concerns |
| Lebov et al. | 2022 | Nicaragua | Lower-middle | Zika fever | Prospective cohort | 126 | 126 | Incidence | Did not pass appropriateness |
| Lebov et al. | 2025 | Nicaragua | Lower-middle | Zika fever | Prospective cohort | 40 | 22 | Incidence | Did not pass appropriateness |
| Mathew et al. | 2011 | India | Lower-middle | Chikungunya | Prospective cohort | 1396 | 3881 | Incidence | Did not pass appropriateness |
| Chu et al | 2025 | Taiwan | High | Dengue fever | Prospective cohort | 816 | 8160 | Hazard ratio | Some concerns |
| Lim et al | 2021 | Singapore | High | Dengue fever | Retrospective cohort | 55870 | 3072309 | Odds ratio | Low |
| Chen et al | 2025 | Taiwan | High | Dengue fever | Case-control | 74422 | 297688 | Odds ratio | Low |
| Gunathilaka et al | 2018 | Sri Lanka | Lower-middle | Dengue fever | Case-control | 53 | 53 | Odds ratio | Some concerns |
| SH Chang et al | 2021 | Taiwan | High | Dengue fever | Retrospective cohort | 398 | 1592 | Hazard ratio | Low |
| Lin et al | 2024 | Taiwan | High | Dengue fever | Retrospective cohort | 48884 | 195536 | Hazard ratio | Some concerns |
| Chien et al | 2020 | Taiwan | High | Dengue fever | Retrospective cohort | 12573 | 62865 | Hazard ratio | Some concerns |
| CC Chang et al | 2021 | Taiwan | High | Dengue fever | Retrospective cohort | 29365 | 117460 | Hazard ratio | High |
| Henderson et al | 2005 | England | High | RSV | Prospective cohort | 150 | 13912 | Hazard ratio | High |
| Coutts et al | 2020 | Scotland | High | RSV | Retrospective cohort | 15795 | 724623 | Odds ratio | Some concerns |
| Stensballe et al | 2009 | Denmark | High | RSV | Retrospective cohort | 848 | 17766 | Relative risk | Did not pass appropriateness |
| Sigurs et al | 1995 | Sweden | High | RSV | Prospective cohort | 47 | 93 | Relative risk | High |
| Sigurs et al | 2000 | Sweden | High | RSV | Prospective cohort | 47 | 93 | Odds ratio | High |
| Sigurs et al | 2005 | Sweden | High | RSV | Prospective cohort | 47 | 93 | Relative risk | High |
| Sigurs et al | 2010 | Sweden | High | RSV | Prospective cohort | 47 | 93 | Odds ratio | High |
| Homaira et al | 2017 | Australia | High | RSV | Retrospective cohort | 31831 | 815685 | Hazard ratio | Some concerns |
| Ehlmaier et al | 2025 | Austria | High | RSV | Prospective cohort | 216 | 201 | Odds ratio | Low |
| Rosas-Salazar et al | 2024 | USA | High | RSV | Prospective cohort | 944 | 797 | Odds ratio | Low |
| Quinn et al | 2023 | Canada | High | Influenza | Retrospective cohort | 17516 | 26499 | Hazard ratio | High |
| Lee et al | 2022 | South Korea | High | Influenza | Retrospective cohort | 2380696 | 21615 | Incidence rate ratio | High |
| de Havenon et al | 2024 | Multinational | NA | Influenza | Retrospective cohort | 77272 | 77272 | Hazard ratio | Some concerns |
| Tesch et al | 2024 | Germany | High | Influenza | Retrospective cohort | 16264 | 29660 | Incidence rate ratio | Some concerns |
| Lo Re III et al | 2022 | USA | High | Influenza | Retrospective cohort | 8269 | 85637 | Hazard ratio | Some concerns |
| Manimunda et al | 2010 | India | Lower-middle | Chikungunya | Other (descriptive surveillance) | 203 | NA | Incidence | Did not pass appropriateness |
| Gerardin et al | 2008 | France (Reunion) | High | Chikungunya | Prospective cohort | 19 | 38 | Incidence | Did not pass appropriateness |
| Charniga et al | 2021 | Colombia | Upper-middle | Zika fever | Other (descriptive surveillance) | 106000 | NA | Incidence | Did not pass appropriateness |
| Pacheco et al | 2021 | Colombia | Upper-middle | Zika fever | Prospective cohort | 60 | NA | Incidence | Did not pass appropriateness |

### Meta-analysis

Chikungunya and Zika studies only reported incidence and were not eligible for meta-analysis.

### RSV-ARI

Considerable variation existed in the length of follow up, sample sizes, and exposure definitions. In addition to asthma, an individual study found increased risks of having any PAS, cardiovascular, respiratory, and neurological sequelae among those with a history of symptomatic RSV-ARI versus COVID-19 patients.^11^ All other studies compared individuals who did and did not have RSV-ARI diagnoses during infancy.

Meta-analysis of RSV-ARI studies focused on asthma outcomes (Figure 4). All estimates were adjusted odds ratios, and individual studies consistently showed higher odds of asthma following symptomatic RSV infection. The pooled odds ratio was 2.93 (95% CI 2.12–4.06), with substantial heterogeneity (prediction interval 0.97–8.84). Sensitivity analysis excluding high-risk studies produced a pooled OR of 2.26 (1.10–4.63) (Supplementary Materials Figure S3).

**Figure 4:**
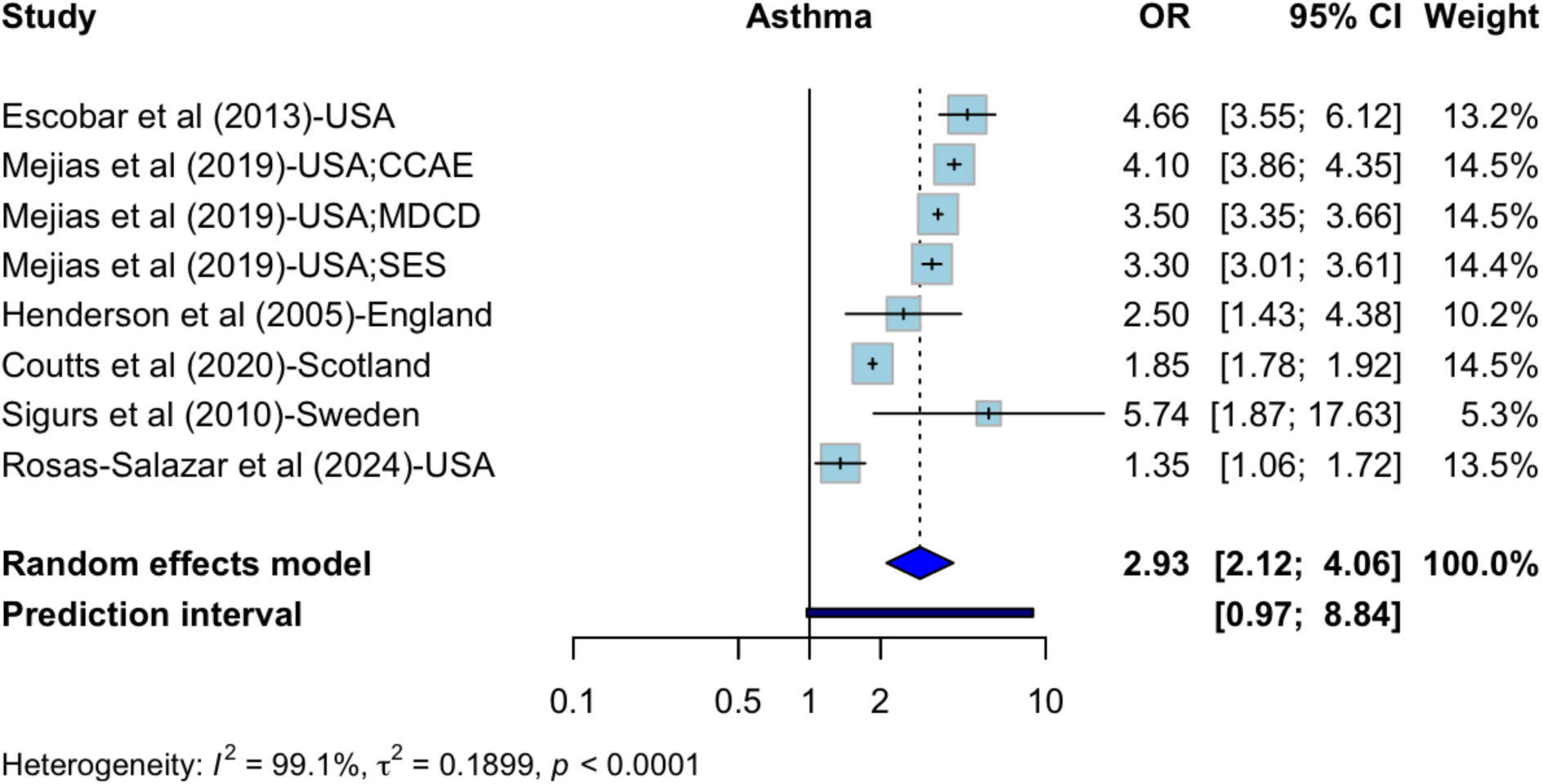
Meta-analysis of post-acute asthmatic outcomes for symptomatic RSV-exposed versus RSV-naïve cohorts.

### Influenza

Individual studies found the influenza group to have lower risks of venous thromboembolism, chronic fatigue syndrome, hair loss, all-cause hospital and emergency department visits and mortality, persistent hypertension, hematological, and endocrine outcomes with COVID-19 patients as the reference group.^49,51–54,58–62^ Conversely, some studies found a significant increase in risk of epilepsy, skin disease, periodontal disease, asthma, Parkinson’s Disease, and, opposite to one of the aforementioned studies, persistent hypertension.^51,55,61^

Substantial variation was observed within time of outcome assessment post-exposure (4 weeks, 31–90 days, 0–180 days) and disease severity (outpatient versus inpatient hospitalization). All studies included in meta-analysis used COVID-19 patients as comparators: three included only patients with the wildtype SARS-CoV-2 strain, of which two incorporated both mild and severe cases,^51,56^, and the other only included hospitalized patients;^61^ another study included wildtype, Alpha, and Delta variants for hospitalized patients;^55^ two studies included the Omicron only, one with hospitalized patients,^58^ and the other with patients diagnosed with an ARI and COVID-19 positive,^52^ and one included all major variants (wildtype, Alpha, Delta, Omicron) for hospitalized patients.^59^

Pooled analyses showed significantly lower risks of neurological outcomes (pooled HR 0.82 [95% CI 0.76–0.89]) and composite outcomes (pooled RR 0.88 [0.82–0.95]) compared with COVID-19 patients. Other pooled outcomes were not statistically significant and showed substantial heterogeneity (Figure 5). Sensitivity analyses excluding high-RoB studies yielded similar results, with cardiovascular outcomes becoming significantly reduced (0.95 [0.93–0.99]) (Supplement Materials Figure S4).

**Figure 5:**
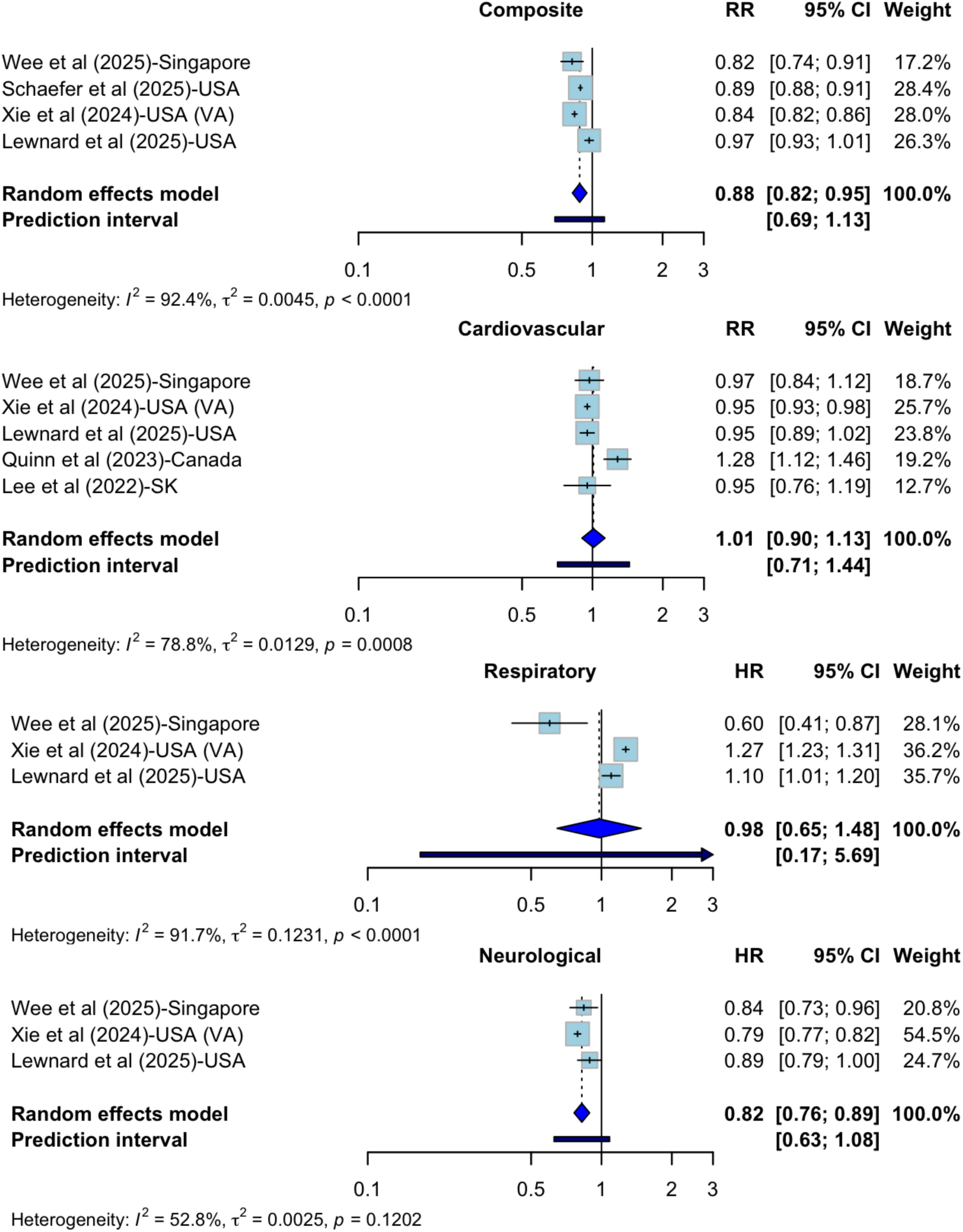
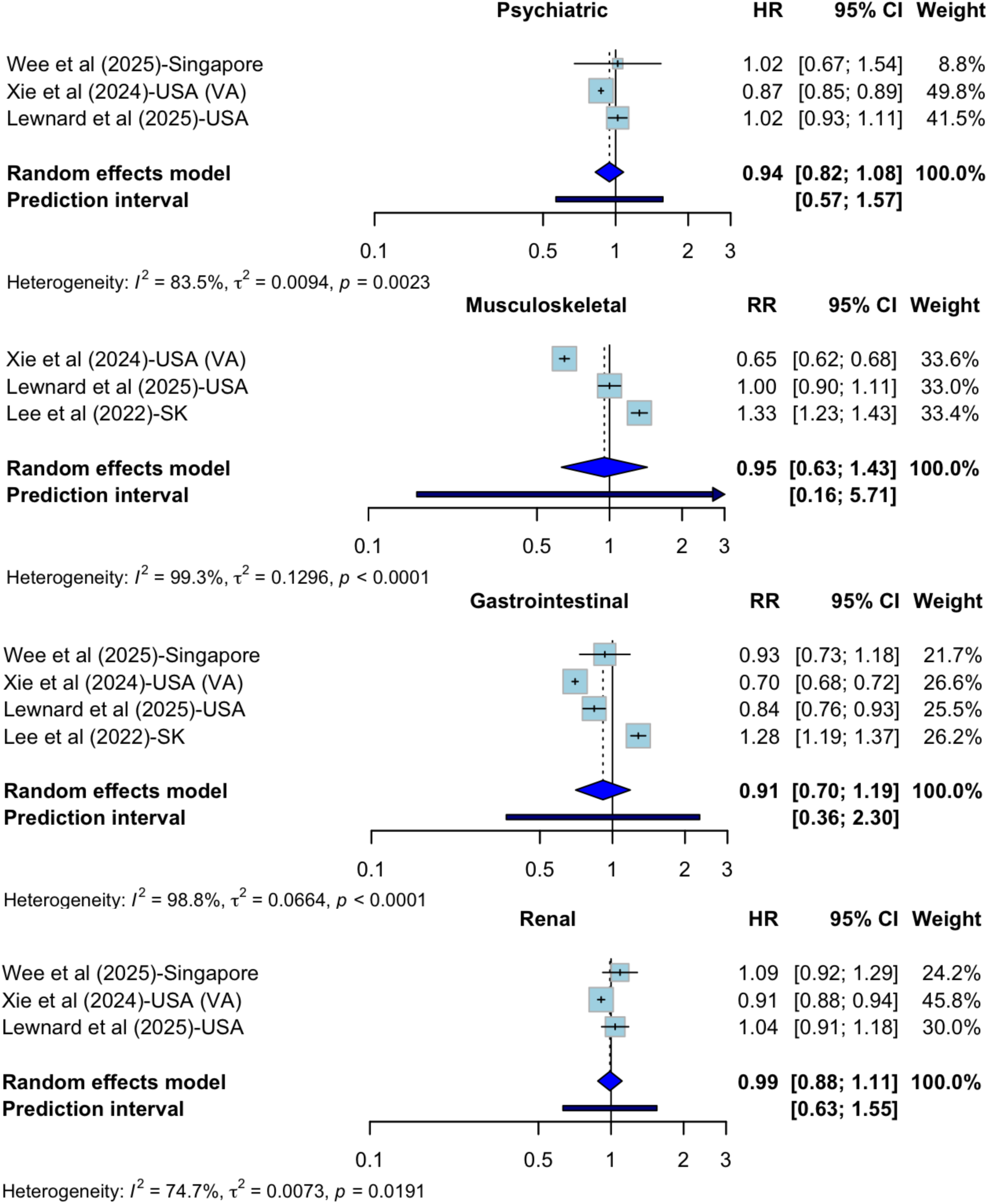
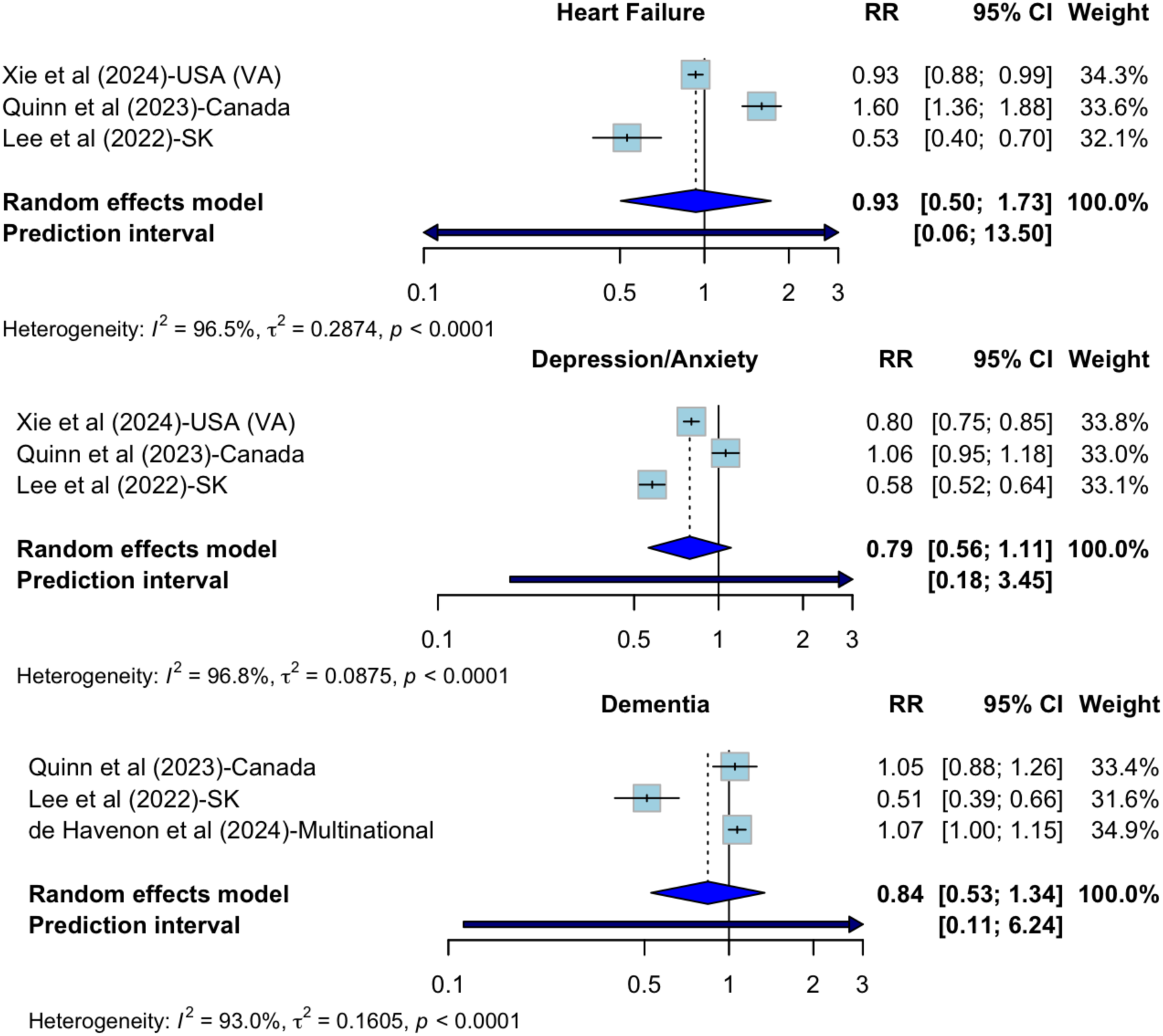
Meta-analysis of post-acute cardiovascular, respiratory, neurological, psychiatric, gastrointestinal, musculoskeletal, renal, and any-sequelae (composite) outcomes, heart failure, depression/anxiety, and dementia for influenza patients versus COVID-19 patients.

### Dengue fever

Variation was less pronounced than for influenza/RSV-ARI studies, except for one study that focused only on a pediatric population (under 18 years), and two studies that used COVID-19-infected individuals as comparators. The remaining studies used any non-dengue individuals as controls. Individual studies found those with a history of symptomatic dengue infection to be significantly associated with lower risk of systemic autoimmune disorders, but higher risks of leukemia, inpatient hospitalization, neurological, and gastrointestinal outcomes.^21,35,39,43–45^

The pediatric cohort using COVID-19 controls yielded non-significant results, but most other studies reported significant associations. Pooled analyses showed increased risks of anxiety (HR: 1.34 [1.01–1.78]), dementia (1.61 [1.10–2.35]), autoimmune (RR: 1.39 [1.17–1.67]), cardiovascular (1.51 [1.27–1.80]), psychiatric (1.17 [1.07–1.28]), and composite post-acute sequelae (1.19 [1.13–1.25]) compared with non-dengue controls (Figure 6). No sensitivity analyses were performed for dengue fever studies, as none had high RoB.

**Figure 6.**
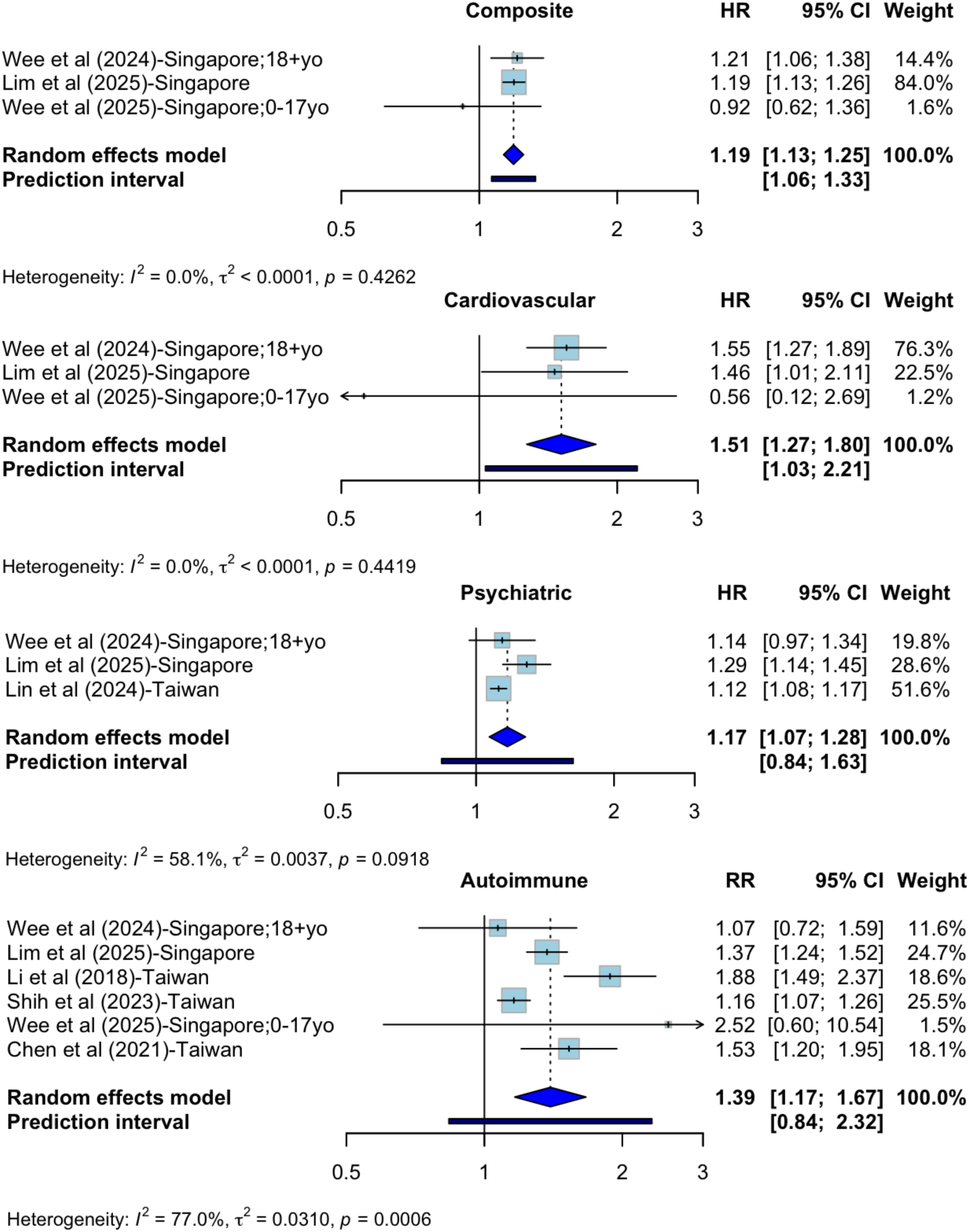
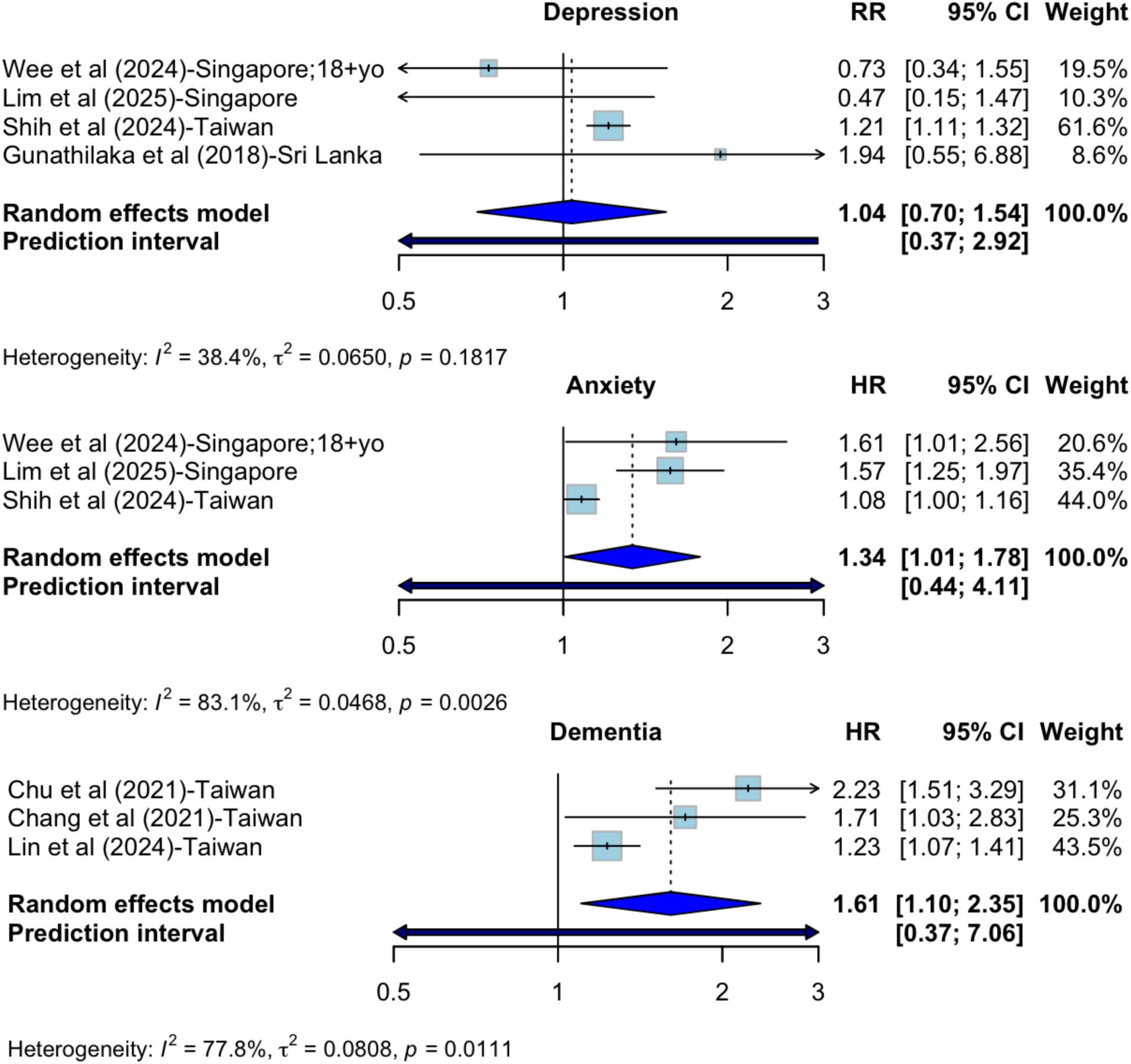
Meta-analysis of post-acute cardiovascular, psychiatric, autoimmune, and any sequelae (composite) outcomes, depression, anxiety, and dementia for dengue fever cases versus dengue-naïve cohorts.

### Publication Bias

Funnel plots were visually inspected to assess potential publication bias. Overall, most plots approximated the expected symmetrical inverted funnel shape. However, the influenza-respiratory sequelae and RSV-asthma exposure-outcome estimates deviated from this pattern. In these two cases, studies with higher precision, represented by larger values of 1/SE, were not clustered near the apex of the funnel around the pooled effect estimate. Instead, they were distributed on both sides of the center, rather than narrowing toward it as would typically be expected for highly precise estimates (Supplementary Materials Figure S5–7).

## Discussion

To our knowledge, this is the most comprehensive synthesis as of 2026 of PAS following major non-COVID-19 respiratory diseases and *Aedes*-borne viruses. We evaluated long-term risks following RSV-ARI, influenza, and dengue fever, while identifying major evidence gaps for chikungunya, Zika, and yellow fever. The limited evidence for these latter diseases likely reflects sporadic transmission patterns and limited availability of longitudinal clinical data in endemic regions, many of which are lower- and middle-income countries (LMICs). All identified studies on Zika and chikungunya were descriptive surveillance or prospective cohort studies, highlighting the need for retrospective cohort analyses in these understudied populations.

Overall, our findings suggest that PAS are not unique to COVID-19 but represent a broader post-viral phenomenon that varies by pathogen and organ system. For RSV-ARI, we observed elevated odds of physician-diagnosed asthma following infection (OR, 2.93 (2.12–4.06). Despite high heterogeneity and observed publication bias, the direction of association was consistent across studies, and exposure definitions were similar, focusing on diagnosed RSV-ARI during infancy. The elevated risks of asthma may be explained via a few different pathways, such as the association between severe RSV diseases in infancy and increased nasal plasmacytoid dendritic cells, which could suggest a raised antiviral response in the airway resulting in the development of asthma,^82^ or a type-2 and type-17 immune response to RSV infection which is associated with recurrent wheezing later, suggesting that differences in immune response may be responsible for the link between early-life RSV infection and future respiratory conditions.^83^ In addition, RSV encodes non-structural proteins that actively suppress type I interferon signaling pathways, impairing early antiviral responses and altering downstream immune regulation, which may contribute to persistent immune dysregulation and increased susceptibility to chronic airway inflammation following early-life infection.^84,85^

In contrast, pooled estimates for influenza, using COVID-19-infected patients as the control group, did not demonstrate increased risk of most organ-specific outcomes. Many included studies were designed to evaluate PAS of COVID-19, with influenza serving as a comparator, which may attenuate observed differences due to more intensive surveillance of COVID-19 patients. Widespread vaccination against both infections may have further reduced disease severity and subsequent risk of PAS. Influenza was significantly associated with lower risks of neurological, cardiovascular, and composite outcomes compared with COVID-19; however, these findings likely underestimate the absolute burden of post-influenza sequelae. Therefore, results should be interpreted as comparative rather than absolute risks.

For dengue fever, pooled analyses demonstrated elevated risks of cardiovascular, autoimmune, psychiatric, anxiety, dementia, and composite post-acute outcomes compared with dengue-naïve controls. These findings are notable given that dengue is often conceptualized as an acute febrile illness despite increasing recognition of prolonged post-acute symptoms. Our results suggest that dengue infection may confer measurable long-term risks across multiple organ systems, consistent with its known multisystem inflammatory effects. NS1-mediated glycocalyx disruption and cytokine-driven capillary leakage may contribute to downstream cardiovascular complications,^86^ while viral disruption of the blood-brain barrier and sustained microglial activation with pro-inflammatory cytokine release driving neuronal damage have been proposed as mechanisms for longer-term psychiatric/cognitive manifestations.^87,88^ Autoimmune sequelae may similarly reflect persistent or dysregulated immune responses following infection.^89^ Disturbances in glucose homeostasis after dengue fever might arise from pancreatic involvement,^90^ cytokine-mediated β-cell dysfunction,^91^ and dysregulation of neuroendocrine stress pathways.^92^ Gastrointestinal and renal manifestations may likewise result from post-infectious immune activation, inflammatory tissue injury,^93^ immune complex deposition, rhabdomyolysis, multiorgan dysfunction, or direct viral effects,^94^ although the evidence remains limited.^95^ While the exact mechanism of pathogenesis for PAS of dengue fever remains unclear, emerging evidence indicates that infection can induce reprogramming of innate immune cells (myeloid-derived cells) similar to observations in COVID-19 and influenza, which can lead to subsequent low-level but sustained inflammation and persist for several years.^96–99^ Elucidating these changes may help explain the risk and spectrum of PAS across multiple organ systems after dengue fever.

Across all studies reviewed, those assessed as low RoB generally included robust adjustment for confounding and clear reporting of follow-up and outcomes. Common concerns included insufficient adjustment for post-exposure interventions, differential outcome ascertainment, and incomplete reporting of missing data. Overall, the direction of bias varied across studies, with no consistent pattern across exposures.

Besides differences in RoB, substantial heterogeneity was observed across studies, reflecting differences in follow-up duration, outcome definitions, and analytic approaches. Although restricting analyses to diagnosis-based outcomes and standardized post-acute periods improved comparability, variability remained. Furthermore, the predominant SARS-CoV-2 variants differed across study periods, with some analyses limited to wildtype infections, others including mixed waves of wildtype, Alpha, Delta, and Omicron variants, and two restricted to Omicron-era infections. Studies including hospitalized COVID-19 cases or earlier variants associated with more severe disease may inflate baseline risks in the reference group, thereby exaggerating the apparent protective effect of influenza/dengue fever/RSV-ARI. Conversely, studies restricted to Omicron-era infections, which are generally associated with milder disease, may attenuate observed differences between influenza/dengue fever/RSV-ARI and COVID-19. Such heterogeneity in reference group severity and variant composition likely contributed to variability in effect estimates and should be considered when interpreting pooled risks.

Some limitations should be acknowledged. First and most importantly, the available evidence remains limited. Despite a broad search strategy across illnesses caused by six distinct pathogens and forward citation searching, only 51 studies met inclusion criteria, and meta-analyses were feasible for just RSV diseases, influenza, and dengue fever, and a subset of outcomes. The number of exposures included in this review posed challenges for systematic retrieval, as incorporating multiple pathogens within a single search strategy per database limited our ability to include the full range of pathogen-specific sequelae terms pertaining to each pathogen. As a result, some relevant studies may not have been captured through database searches alone. Additionally, post-acute outcomes are often broadly defined using terms such as “multisystem” or “multiorgan,” without consistent specification of included organ systems, making it difficult to comprehensively capture all relevant outcomes within search strategies. This further contributed to reliance on forward citation searching, which yielded a substantial proportion of included studies. Future systematic reviews may benefit from focusing on individual diseases or related groups of diseases, and specific organ systems or sequelae, allowing for more tailored search strategies that incorporate the most relevant sets of sequelae investigated for each pathogen. No eligible effect estimates were available for yellow fever, and data for chikungunya and Zika fever were insufficient for pooling. This limited data availability restricts the precision of pooled estimates and further research studies would be necessary to explore effect modifications by age, severity of acute illness, vaccination status, or geographic setting. Second, heterogeneity across studies was substantial. Follow-up windows varied, and although we prioritized timepoints closest to a standardized post-acute definition, residual differences likely remain. Outcome definitions were also heterogeneous, with some studies relying on ICD-coded diagnoses for retrospective studies, and others on a variety of tests (antigen, PCR, and others) for the prospective studies, introducing variability in outcome measurement and potential misclassification. Third, RoB was a significant concern, since only a relatively small proportion of studies were assessed as low risk. Many lacked adequate adjustment for confounding, particularly differences in health-seeking behavior, healthcare access, socioeconomic status (which contributes to exposure and outcome reporting), and post-exposure interventions. In some cases, differential outcome ascertainment between exposed and control groups may have biased estimates toward apparent harm, while in others, under-ascertainment of exposure may have biased results toward the null. The overall direction of bias was inconsistent across studies and pathogens, limiting our ability to infer whether pooled estimates systematically overestimate or underestimate true associations.

Despite these limitations, our findings have several implications for public health policy and clinical practice. First, recognition that post-acute sequelae extend beyond COVID-19 underscores the need for broader post-infectious surveillance systems. Health systems could consider integrating increased long-term follow-up for patients recovering from RSV diseases and dengue fever, especially among high-risk populations. Second, the observed associations between symptomatic RSV infection and asthma may inform vaccine cost-effectiveness analyses and prioritization strategies by incorporating potential long-term morbidity, not solely acute incidence and mortality. Third, comparative analyses between viral infections highlight the importance of standardized definitions of post-acute periods and outcome measurement, which would facilitate cross-pathogen comparisons and more reliable burden estimation. Future research should prioritize well-designed, population-based cohort studies with clearly defined exposure ascertainment, standardized post-acute follow-up windows, and consistent outcome definitions. Direct comparisons with uninfected control groups, rather than with other infected populations, would better quantify absolute risk. Greater representation from low- and middle-income countries is also essential, particularly for emerging infectious diseases, which disproportionately affect these regions. Finally, to improve the quality and comparability of reporting on long-term sequelae, studies should ideally use cohort designs comparing exposed versus non-exposed populations, measure outcomes using validated methods such as clinician diagnoses or WHO guidelines rather than self-reported questionnaires, apply standardized outcome definitions (e.g., ICD codes), and follow participants for a standardized duration based on the virus-specific post-acute period.

This systematic review and meta-analysis demonstrates that post-acute sequelae represent a measurable and clinically relevant component of the disease burden for several common viral infections, especially RSV-ARI and dengue fever. However, currently available evidence remains fragmented and limited. Strengthening methodological consistency and expanding high-quality longitudinal data collection will be critical to accurately characterize the long-term consequences of viral infections and inform prevention and follow-up strategies.

## Contributors

LJP contributed to supervision, systematic search, screening, data extraction, risk of bias assessment, meta-analysis, and manuscript writing. BX contributed to data extraction, risk of bias assessment, and manuscript writing. ELWC and JYC contributed to manuscript writing, review and editing. RR and BZML contributed to the initial systematic search and data extraction. ILEW and JTL contributed to conceptualization, supervision, and manuscript review and editing. DCBL, EEO, and KBT contributed to manuscript review and editing. All authors reviewed and approved the final manuscript.

## Data Availability

No data were generated by this study, all data used are available from the original studies reviewed and cited in this systematic review. Data synthesized in this study are available within the manuscript and its supplemental materials.

## Data sharing statement

All data utilized or synthesized to support the findings in this study are available within this manuscript its Supplementary Materials.

## Supplementary Materials

### Appendix A. Full Search Terms Among 5 Databases

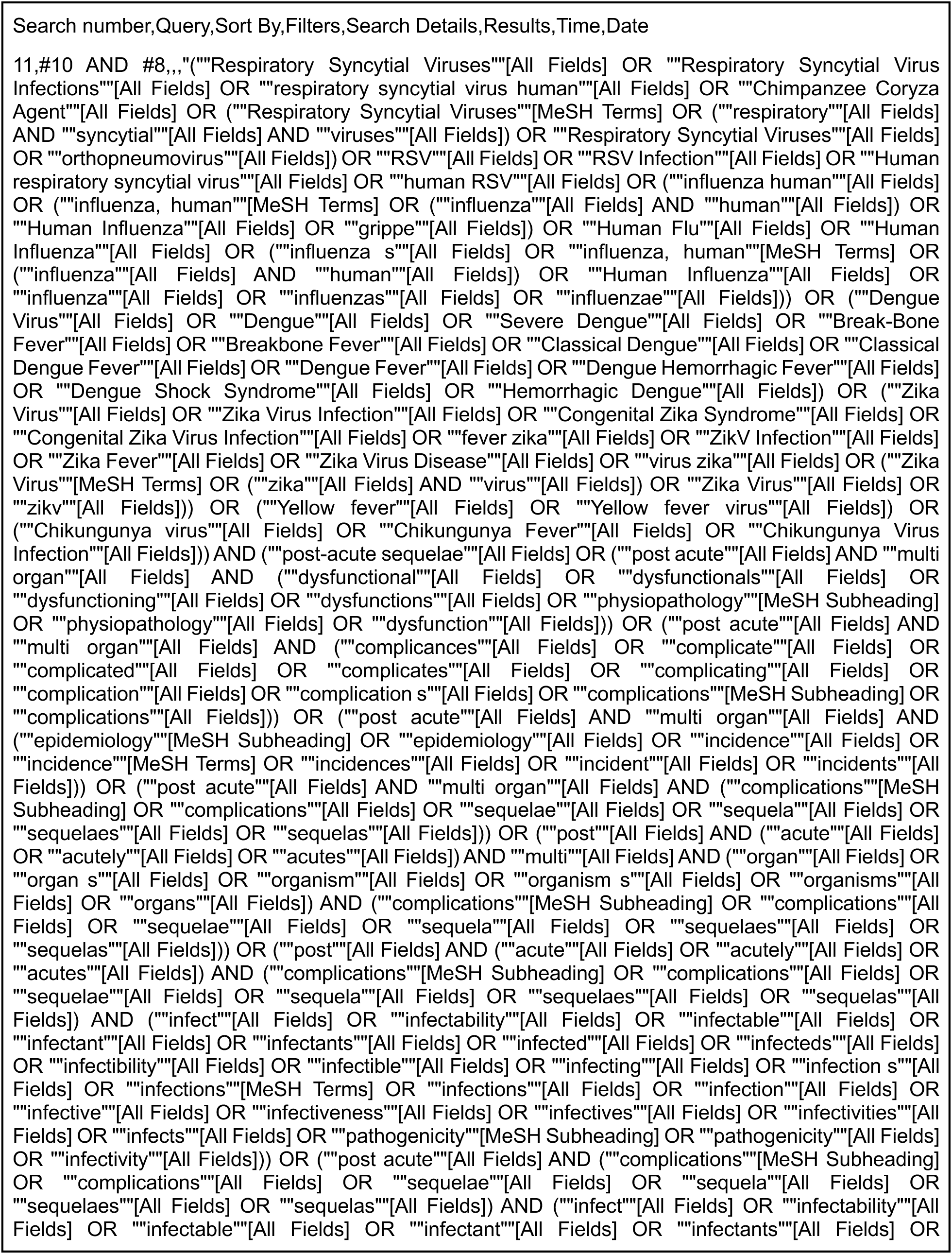
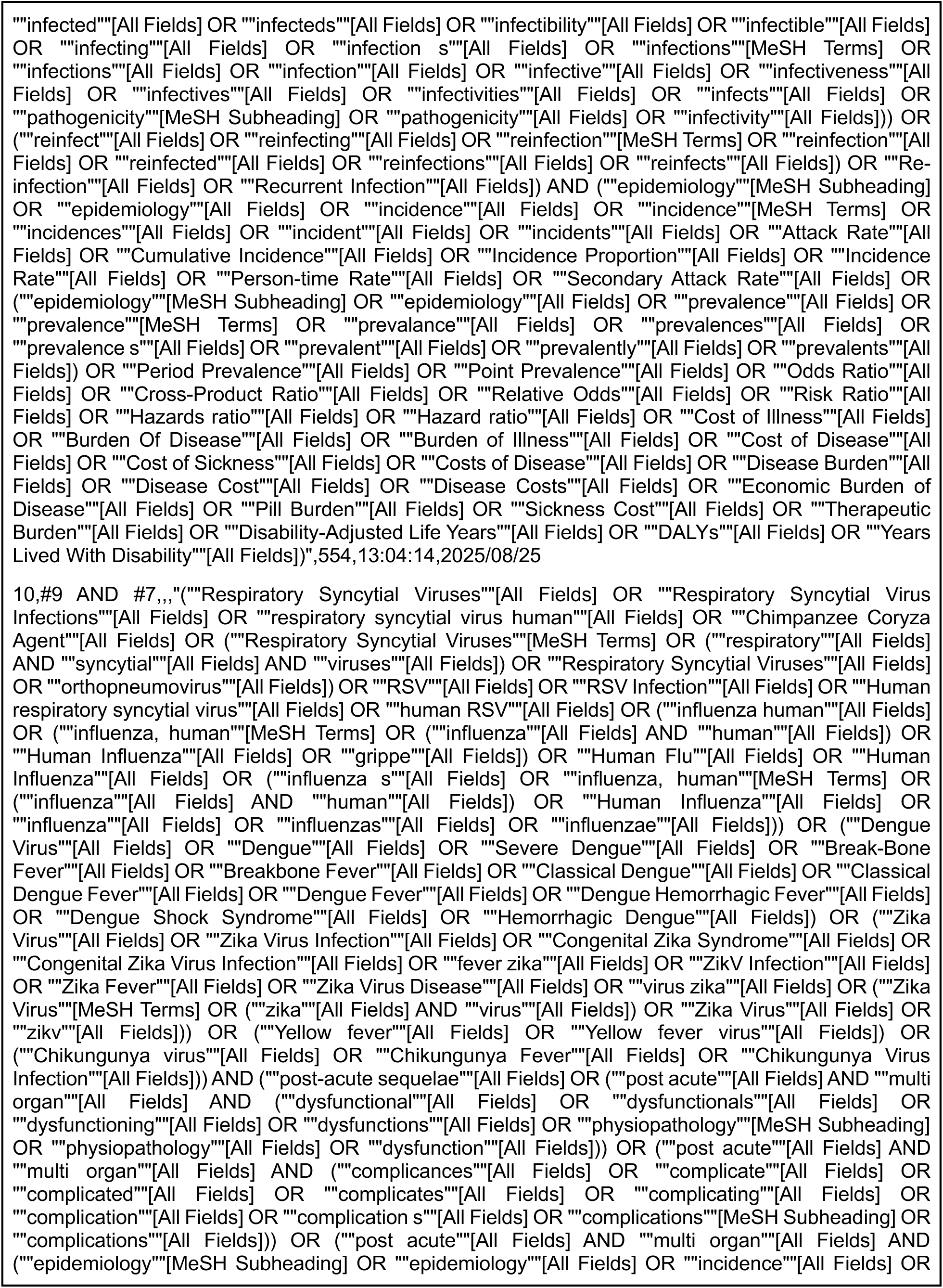
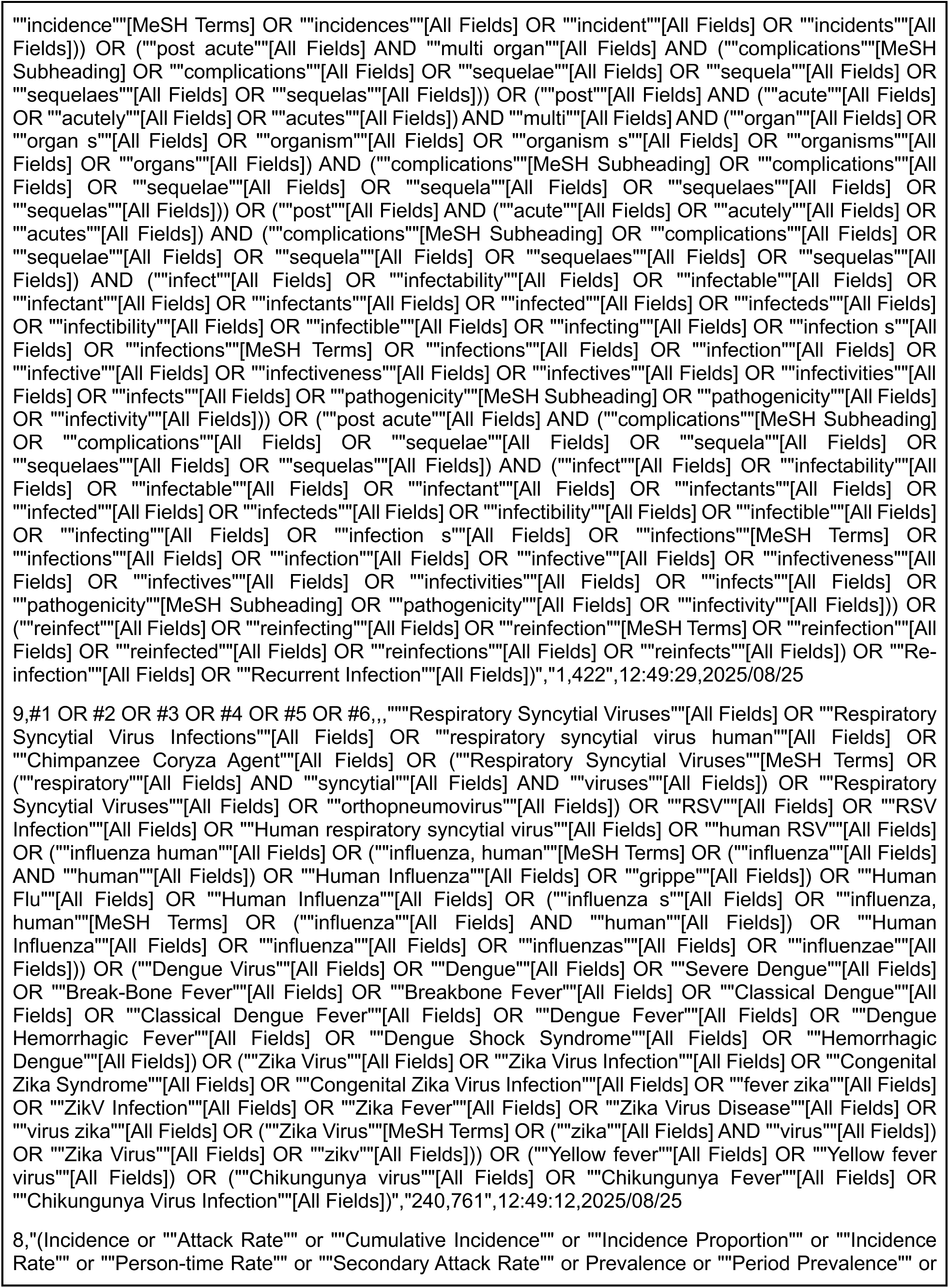
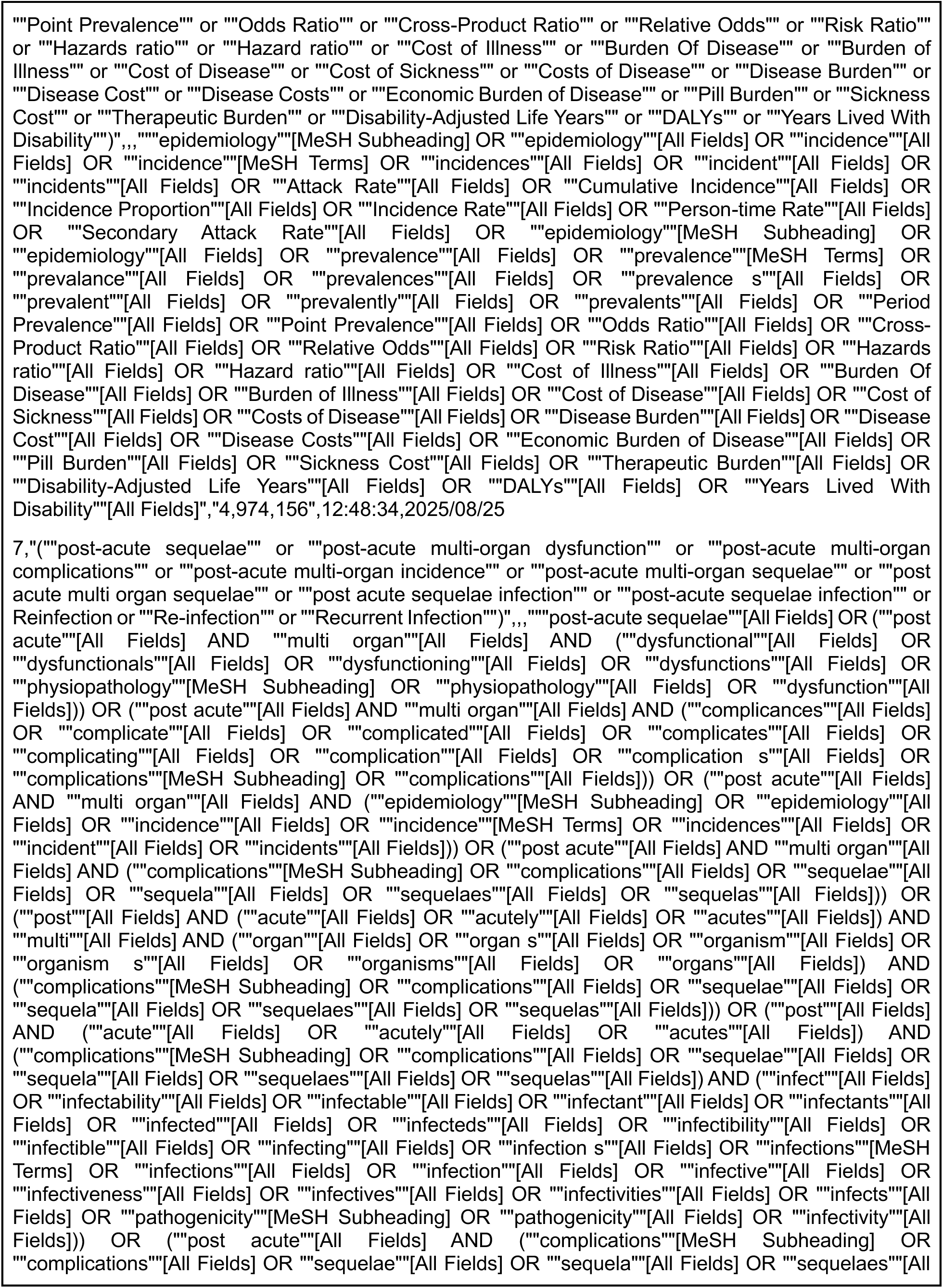
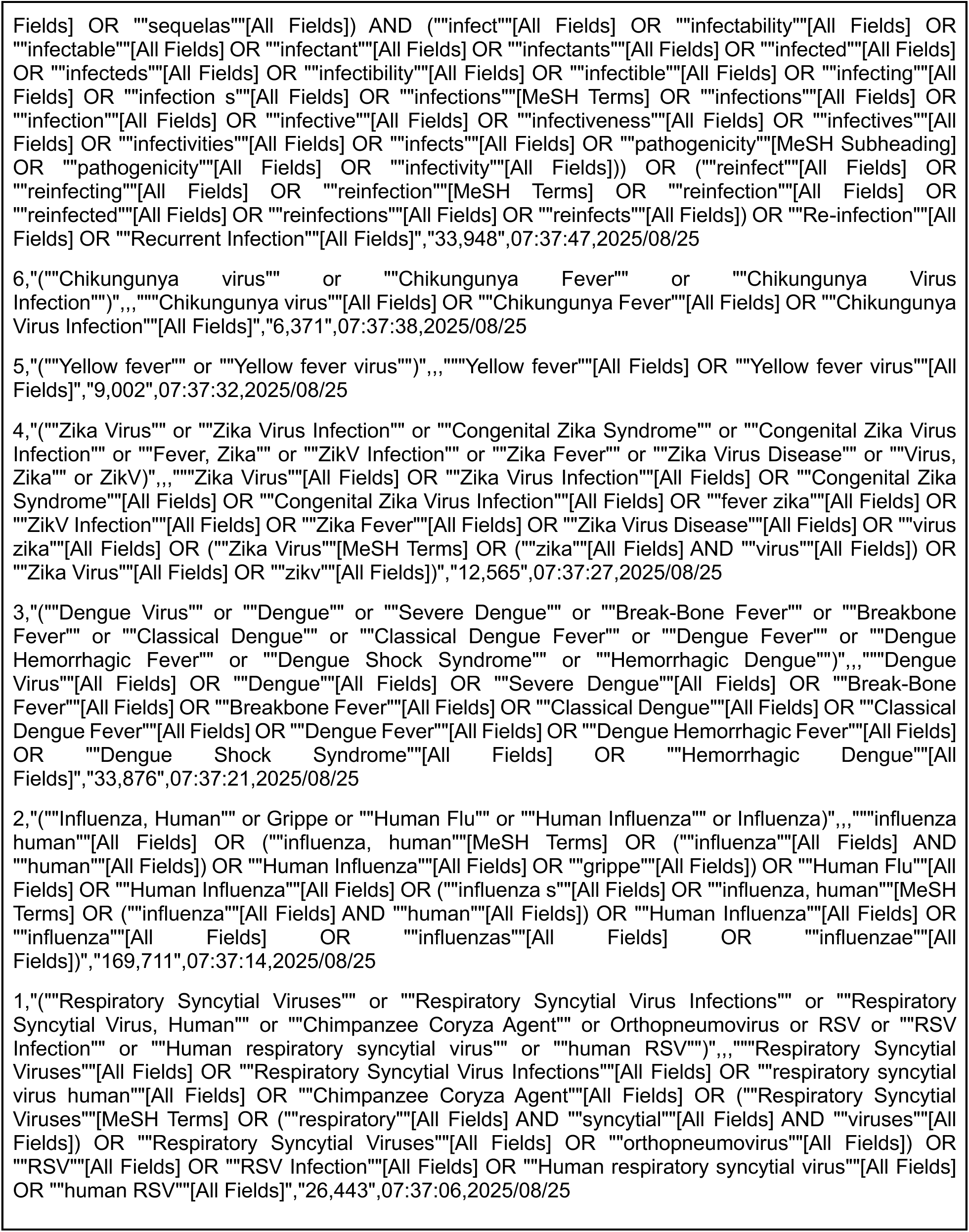
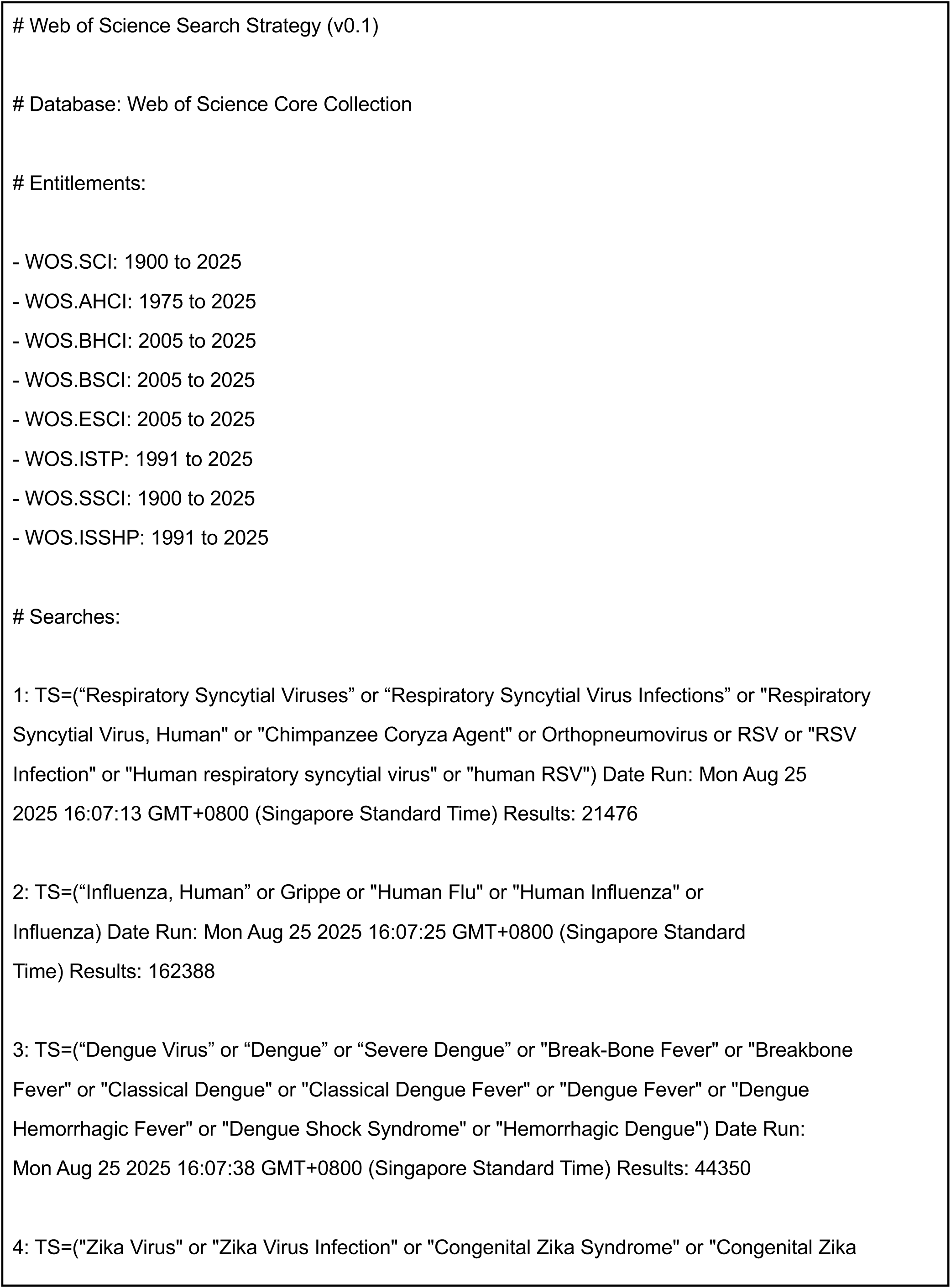
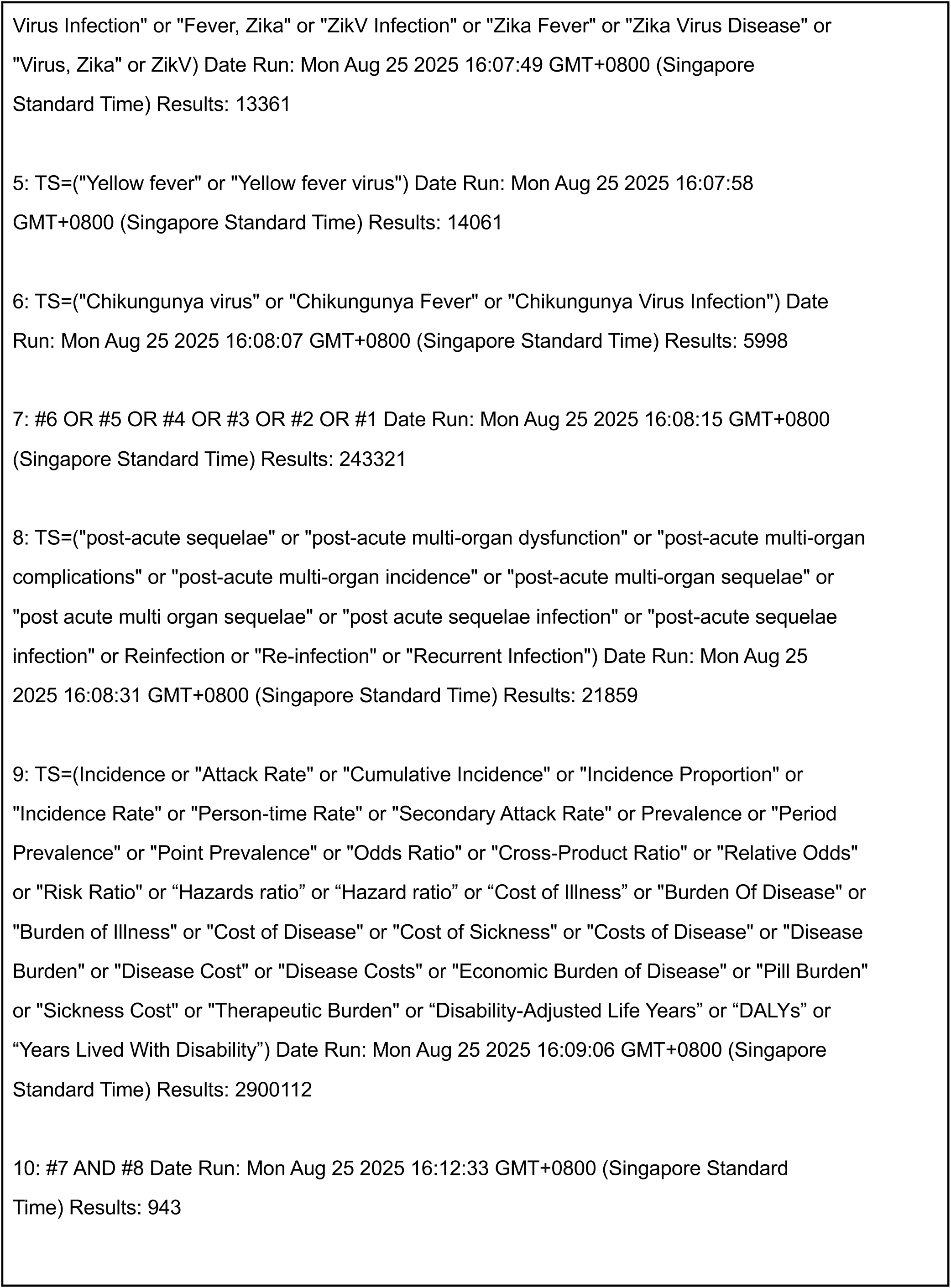
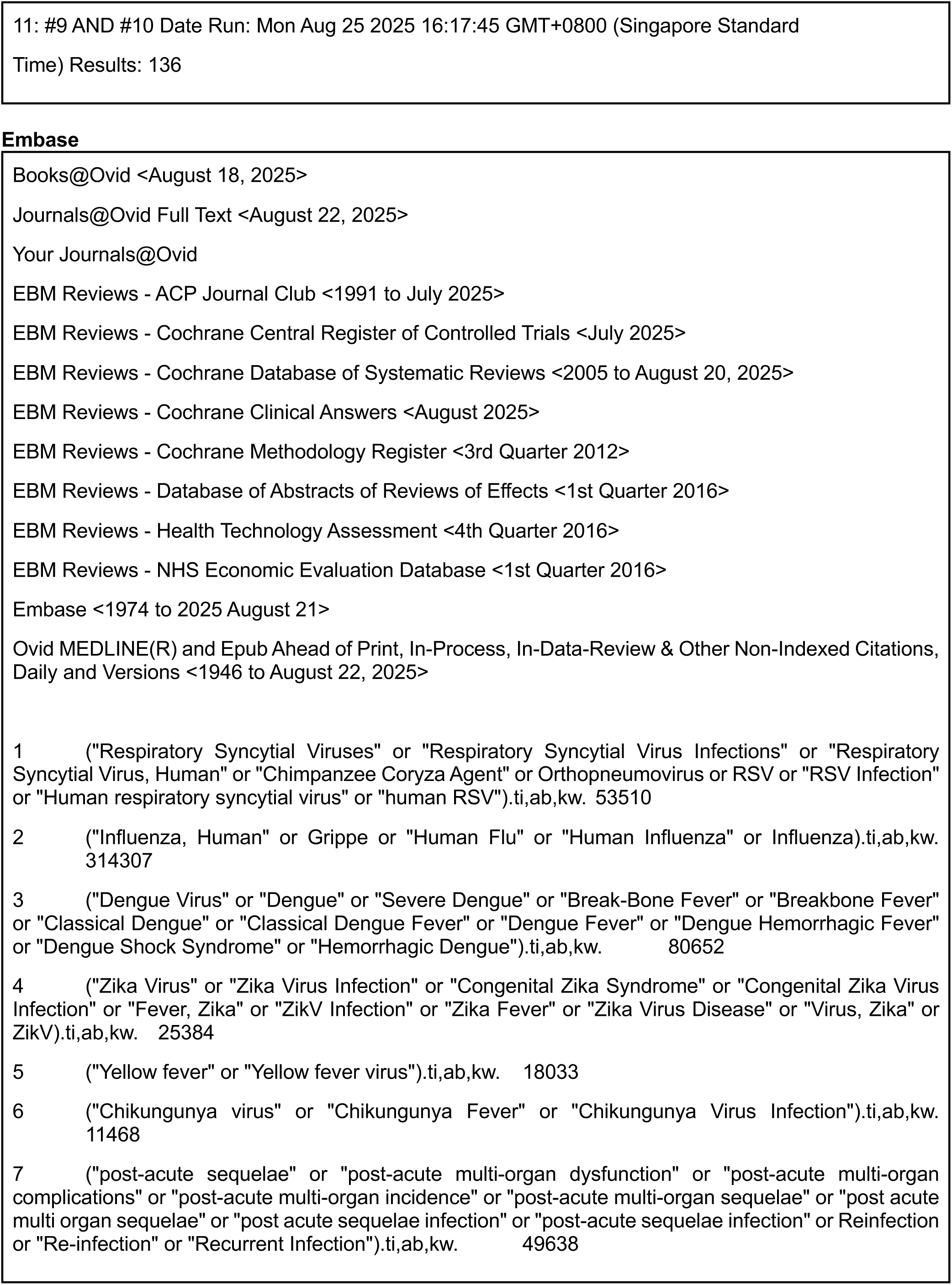
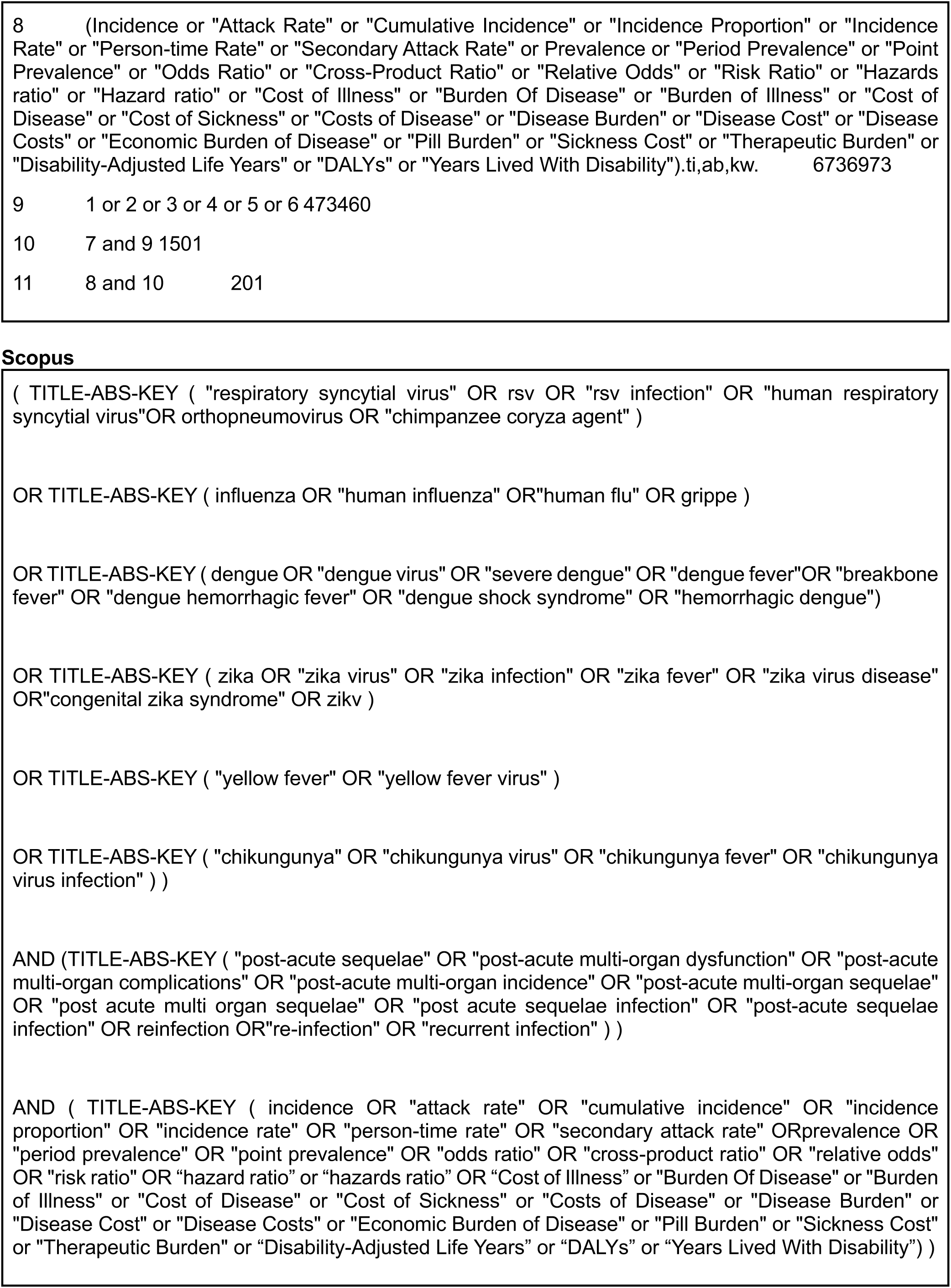
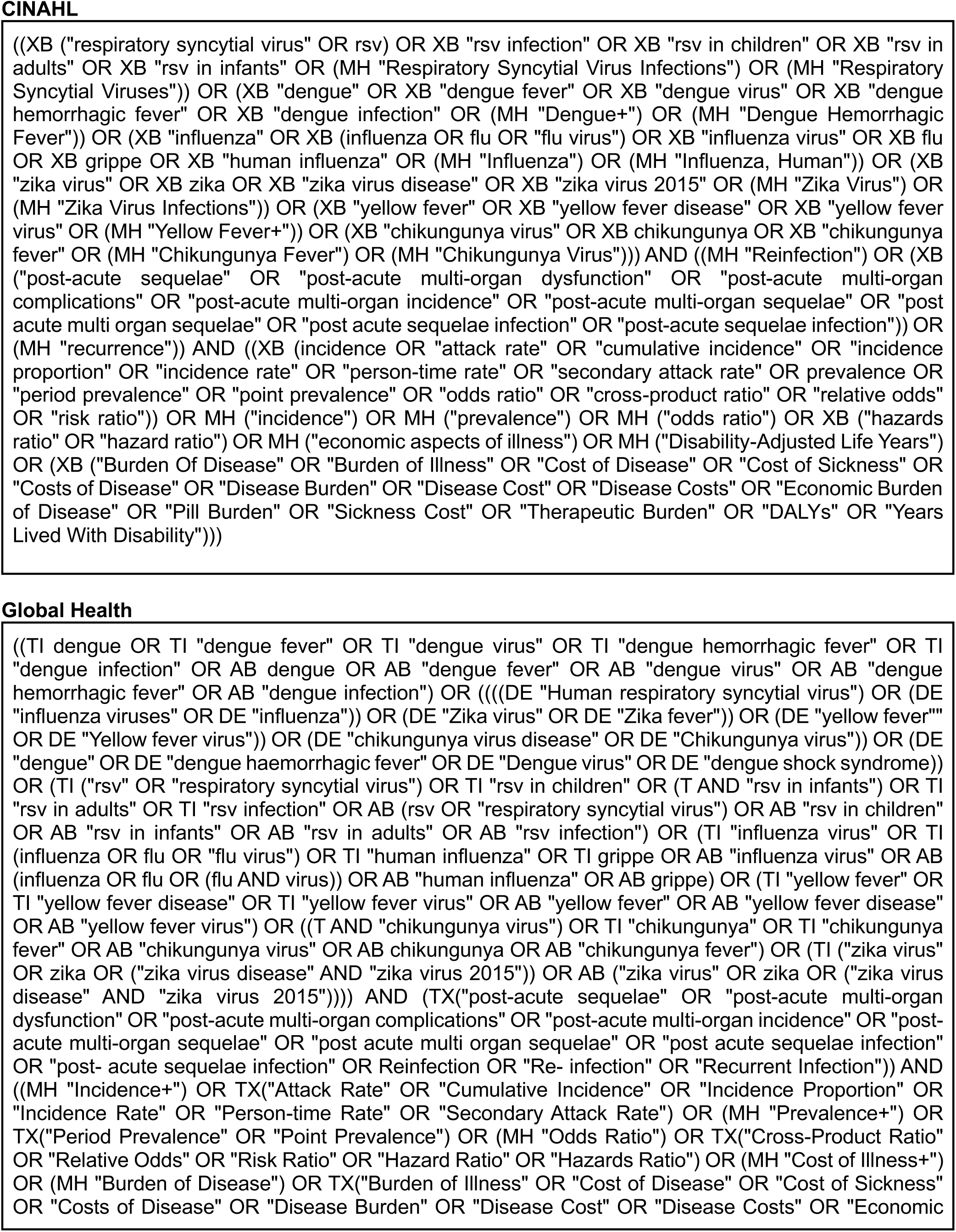
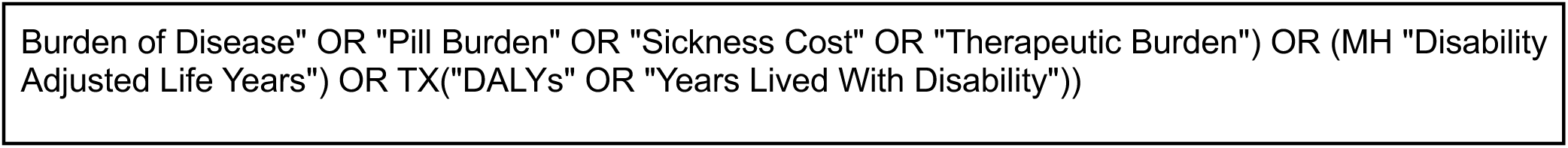

### Appendix B. List of Studies Excluded After Full-text Review

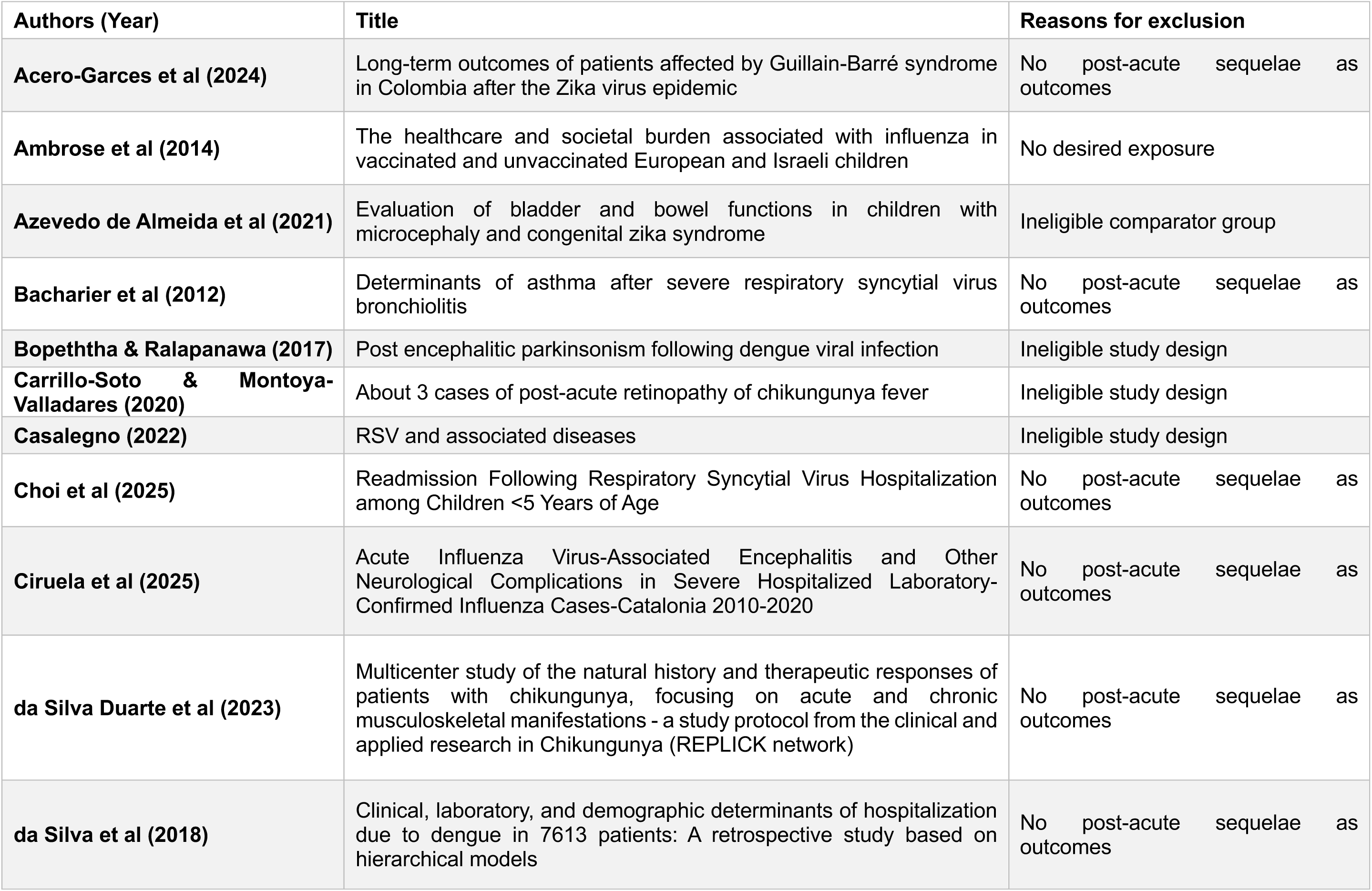
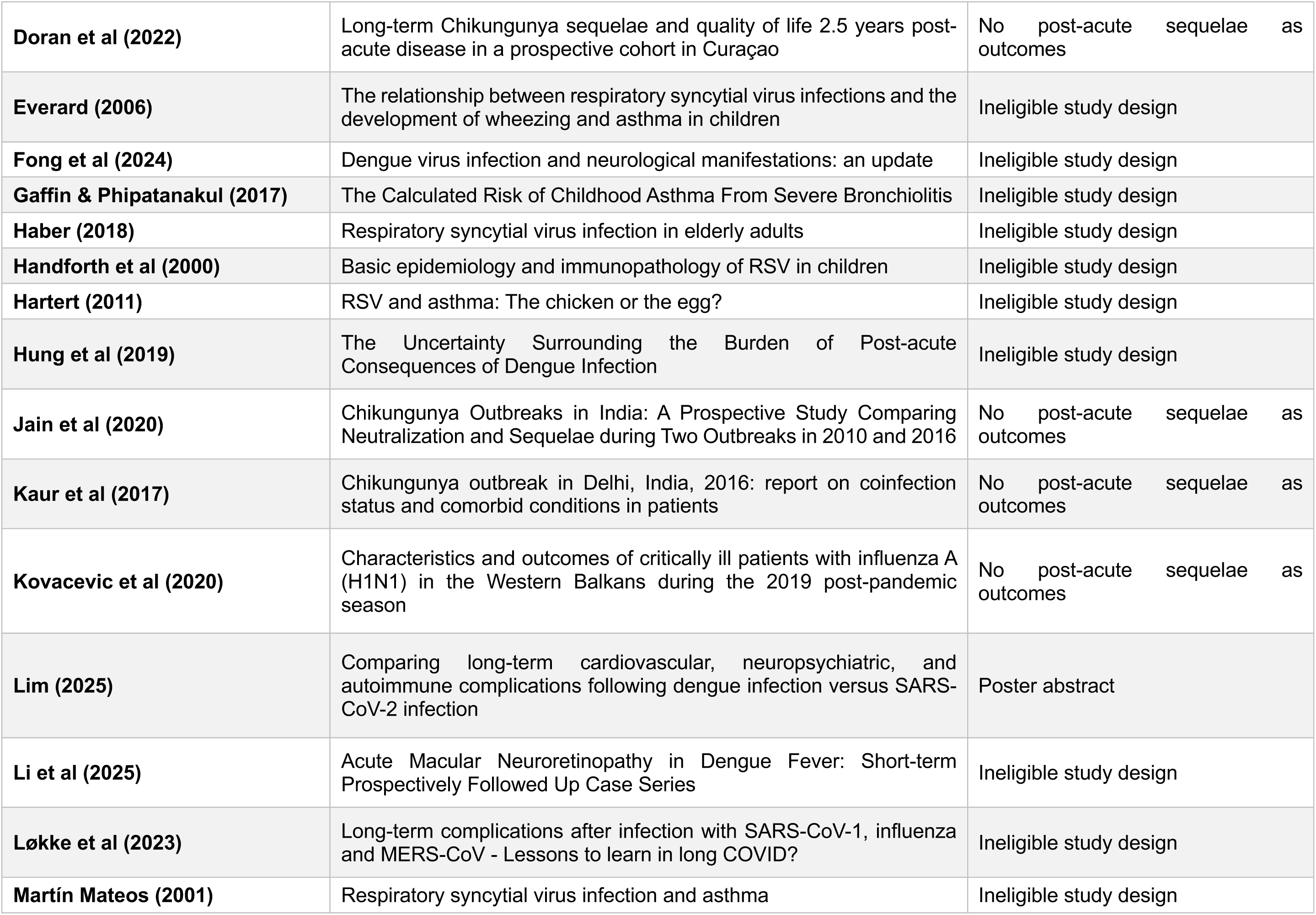
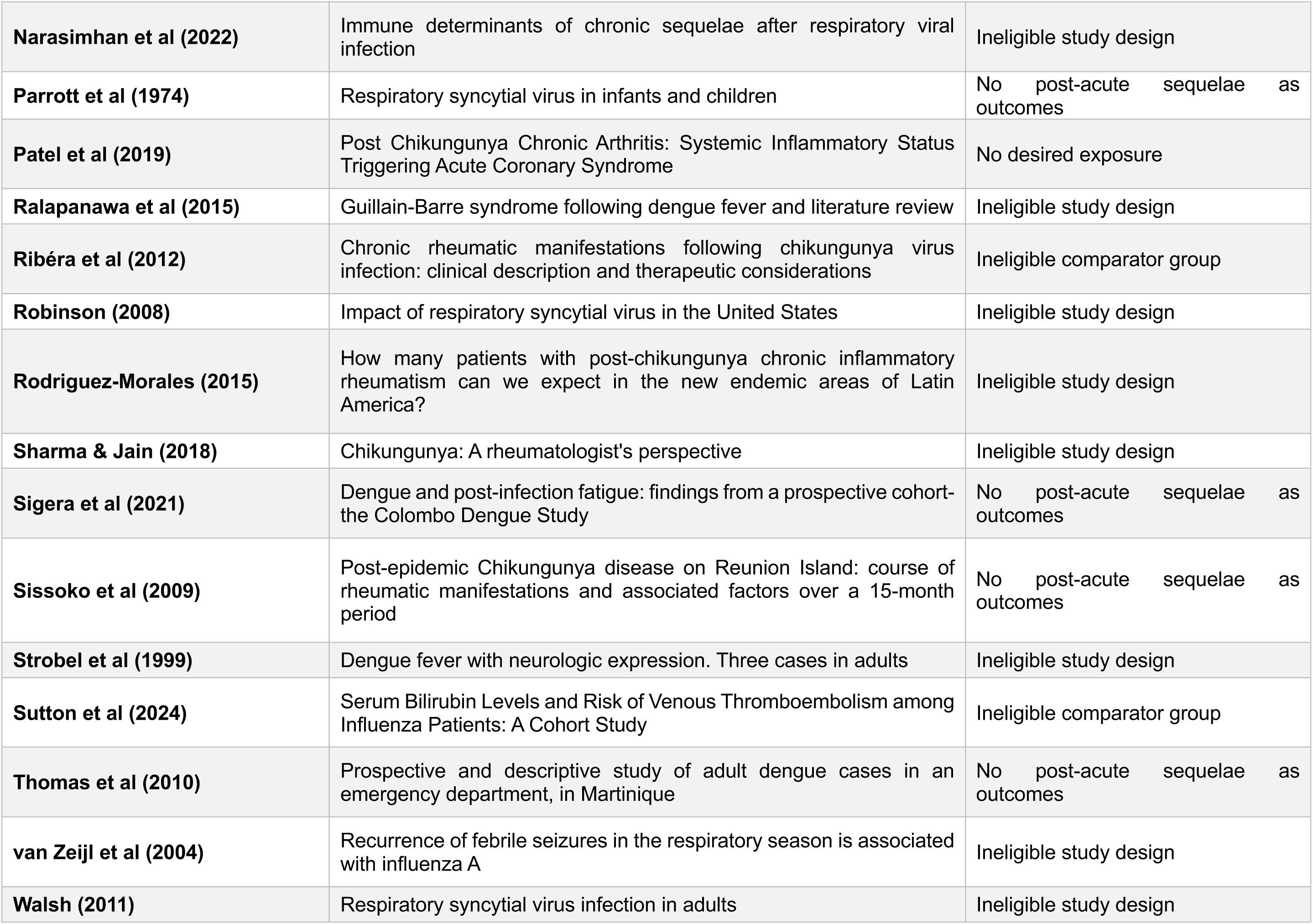
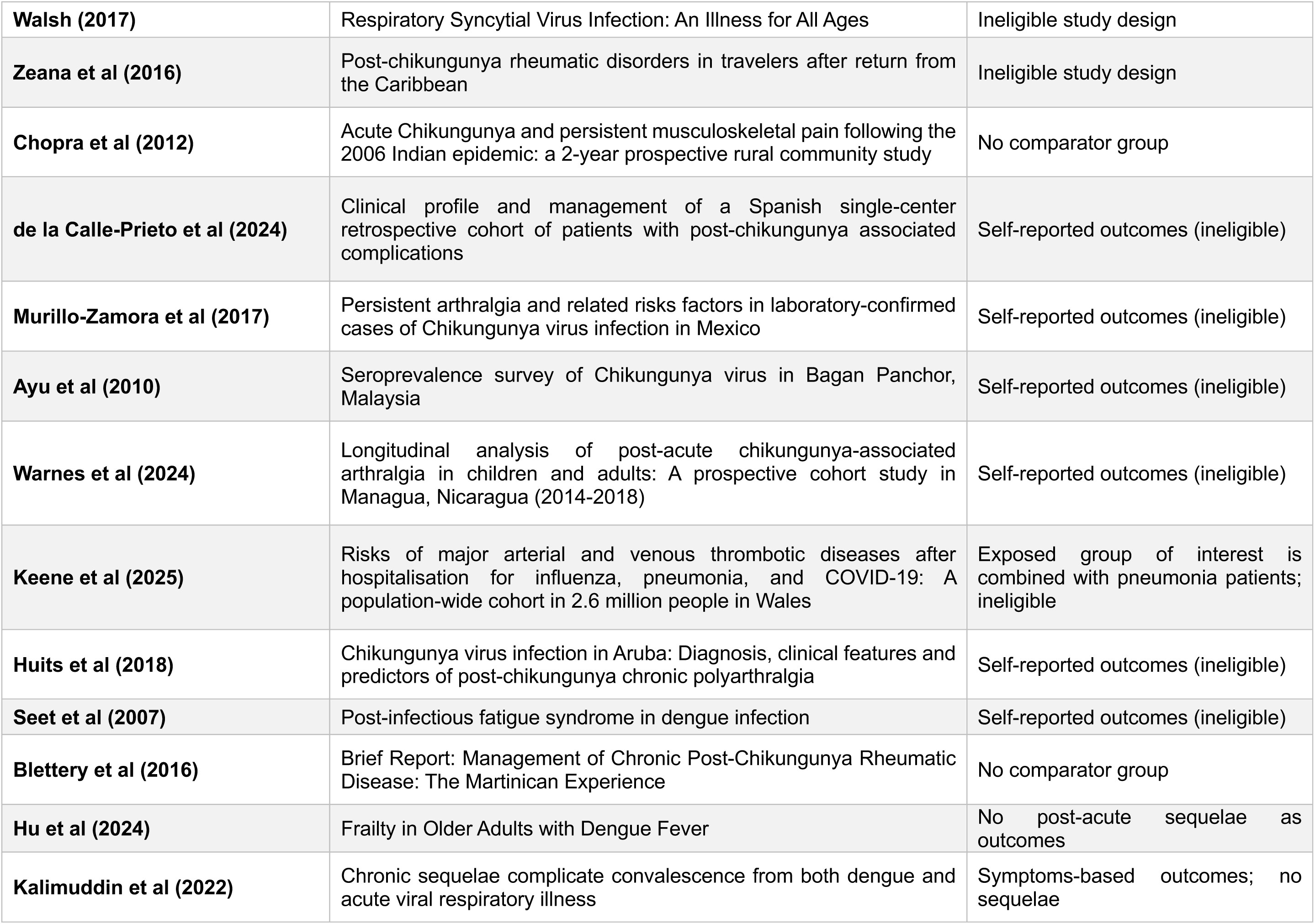
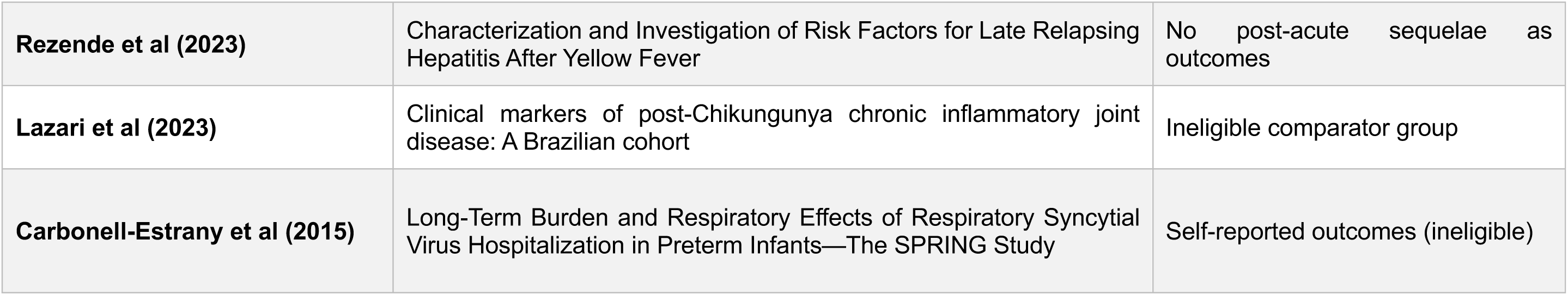

| Authors (Year) | Title | Reasons for exclusion |
| --- | --- | --- |
| **Acero-Garces et al (2024)** | Long-term outcomes of patients affected by Guillain-Barré syndrome in Colombia after the Zika virus epidemic | No post-acute sequelae as outcomes |
| **Ambrose et al (2014)** | The healthcare and societal burden associated with influenza in vaccinated and unvaccinated European and Israeli children | No desired exposure |
| **Azevedo de Almeida et al (2021)** | Evaluation of bladder and bowel functions in children with microcephaly and congenital zika syndrome | Ineligible comparator group |
| **Bacharier et al (2012)** | Determinants of asthma after severe respiratory syncytial virus bronchiolitis | No post-acute sequelae as outcomes |
| **Bopeththa & Ralapanawa (2017)** | Post encephalitic parkinsonism following dengue viral infection | Ineligible study design |
| **Carrillo-Soto & Montoya-Valladares (2020)** | About 3 cases of post-acute retinopathy of chikungunya fever | Ineligible study design |
| **Casalegno (2022)** | RSV and associated diseases | Ineligible study design |
| **Choi et al (2025)** | Readmission Following Respiratory Syncytial Virus Hospitalization among Children <5 Years of Age | No post-acute sequelae as outcomes |
| **Ciruela et al (2025)** | Acute Influenza Virus-Associated Encephalitis and Other Neurological Complications in Severe Hospitalized Laboratory-Confirmed Influenza Cases-Catalonia 2010-2020 | No post-acute sequelae as outcomes |
| **da Silva Duarte et al (2023)** | Multicenter study of the natural history and therapeutic responses of patients with chikungunya, focusing on acute and chronic musculoskeletal manifestations - a study protocol from the clinical and applied research in Chikungunya (REPLICK network) | No post-acute sequelae as outcomes |
| **da Silva et al (2018)** | Clinical, laboratory, and demographic determinants of hospitalization due to dengue in 7613 patients: A retrospective study based on hierarchical models | No post-acute sequelae as outcomes |
| **Doran et al (2022)** | Long-term Chikungunya sequelae and quality of life 2.5 years post-acute disease in a prospective cohort in Curaçao | No post-acute sequelae as outcomes |
| **Everard (2006)** | The relationship between respiratory syncytial virus infections and the development of wheezing and asthma in children | Ineligible study design |
| **Fong et al (2024)** | Dengue virus infection and neurological manifestations: an update | Ineligible study design |
| **Gaffin & Phipatanakul (2017)** | The Calculated Risk of Childhood Asthma From Severe Bronchiolitis | Ineligible study design |
| **Haber (2018)** | Respiratory syncytial virus infection in elderly adults | Ineligible study design |
| **Handforth et al (2000)** | Basic epidemiology and immunopathology of RSV in children | Ineligible study design |
| **Hartert (2011)** | RSV and asthma: The chicken or the egg? | Ineligible study design |
| **Hung et al (2019)** | The Uncertainty Surrounding the Burden of Post-acute Consequences of Dengue Infection | Ineligible study design |
| **Jain et al (2020)** | Chikungunya Outbreaks in India: A Prospective Study Comparing Neutralization and Sequelae during Two Outbreaks in 2010 and 2016 | No post-acute sequelae as outcomes |
| **Kaur et al (2017)** | Chikungunya outbreak in Delhi, India, 2016: report on coinfection status and comorbid conditions in patients | No post-acute sequelae as outcomes |
| **Kovacevic et al (2020)** | Characteristics and outcomes of critically ill patients with influenza A (H1N1) in the Western Balkans during the 2019 post-pandemic season | No post-acute sequelae as outcomes |
| **Lim (2025)** | Comparing long-term cardiovascular, neuropsychiatric, and autoimmune complications following dengue infection versus SARS-CoV-2 infection | Poster abstract |
| **Li et al (2025)** | Acute Macular Neuroretinopathy in Dengue Fever: Short-term Prospectively Followed Up Case Series | Ineligible study design |
| **Løkke et al (2023)** | Long-term complications after infection with SARS-CoV-1, influenza and MERS-CoV - Lessons to learn in long COVID? | Ineligible study design |
| **Martín Mateos (2001)** | Respiratory syncytial virus infection and asthma | Ineligible study design |
| **Narasimhan et al (2022)** | Immune determinants of chronic sequelae after respiratory viral infection | Ineligible study design |
| **Parrott et al (1974)** | Respiratory syncytial virus in infants and children | No post-acute sequelae as outcomes |
| **Patel et al (2019)** | Post Chikungunya Chronic Arthritis: Systemic Inflammatory Status Triggering Acute Coronary Syndrome | No desired exposure |
| **Ralapanawa et al (2015)** | Guillain-Barre syndrome following dengue fever and literature review | Ineligible study design |
| **Ribéra et al (2012)** | Chronic rheumatic manifestations following chikungunya virus infection: clinical description and therapeutic considerations | Ineligible comparator group |
| **Robinson (2008)** | Impact of respiratory syncytial virus in the United States | Ineligible study design |
| **Rodriguez-Morales (2015)** | How many patients with post-chikungunya chronic inflammatory rheumatism can we expect in the new endemic areas of Latin America? | Ineligible study design |
| **Sharma & Jain (2018)** | Chikungunya: A rheumatologist's perspective | Ineligible study design |
| **Sigera et al (2021)** | Dengue and post-infection fatigue: findings from a prospective cohort-the Colombo Dengue Study | No post-acute sequelae as outcomes |
| **Sissoko et al (2009)** | Post-epidemic Chikungunya disease on Reunion Island: course of rheumatic manifestations and associated factors over a 15-month period | No post-acute sequelae as outcomes |
| **Strobel et al (1999)** | Dengue fever with neurologic expression. Three cases in adults | Ineligible study design |
| **Sutton et al (2024)** | Serum Bilirubin Levels and Risk of Venous Thromboembolism among Influenza Patients: A Cohort Study | Ineligible comparator group |
| **Thomas et al (2010)** | Prospective and descriptive study of adult dengue cases in an emergency department, in Martinique | No post-acute sequelae as outcomes |
| **van Zeijl et al (2004)** | Recurrence of febrile seizures in the respiratory season is associated with influenza A | Ineligible study design |
| **Walsh (2011)** | Respiratory syncytial virus infection in adults | Ineligible study design |
| **Walsh (2017)** | Respiratory Syncytial Virus Infection: An Illness for All Ages | Ineligible study design |
| **Zeana et al (2016)** | Post-chikungunya rheumatic disorders in travelers after return from the Caribbean | Ineligible study design |
| **Chopra et al (2012)** | Acute Chikungunya and persistent musculoskeletal pain following the 2006 Indian epidemic: a 2-year prospective rural community study | No comparator group |
| **de la Calle-Prieto et al (2024)** | Clinical profile and management of a Spanish single-center retrospective cohort of patients with post-chikungunya associated complications | Self-reported outcomes (ineligible) |
| **Murillo-Zamora et al (2017)** | Persistent arthralgia and related risks factors in laboratory-confirmed cases of Chikungunya virus infection in Mexico | Self-reported outcomes (ineligible) |
| **Ayu et al (2010)** | Seroprevalence survey of Chikungunya virus in Bagan Panchor, Malaysia | Self-reported outcomes (ineligible) |
| **Warnes et al (2024)** | Longitudinal analysis of post-acute chikungunya-associated arthralgia in children and adults: A prospective cohort study in Managua, Nicaragua (2014-2018) | Self-reported outcomes (ineligible) |
| **Keene et al (2025)** | Risks of major arterial and venous thrombotic diseases after hospitalisation for influenza, pneumonia, and COVID-19: A population-wide cohort in 2.6 million people in Wales | Exposed group of interest is combined with pneumonia patients; ineligible |
| **Huits et al (2018)** | Chikungunya virus infection in Aruba: Diagnosis, clinical features and predictors of post-chikungunya chronic polyarthralgia | Self-reported outcomes (ineligible) |
| **Seet et al (2007)** | Post-infectious fatigue syndrome in dengue infection | Self-reported outcomes (ineligible) |
| **Blettery et al (2016)** | Brief Report: Management of Chronic Post-Chikungunya Rheumatic Disease: The Martinican Experience | No comparator group |
| **Hu et al (2024)** | Frailty in Older Adults with Dengue Fever | No post-acute sequelae as outcomes |
| **Kalimuddin et al (2022)** | Chronic sequelae complicate convalescence from both dengue and acute viral respiratory illness | Symptoms-based outcomes; no sequelae |
| **Rezende et al (2023)** | Characterization and Investigation of Risk Factors for Late Relapsing Hepatitis After Yellow Fever | No post-acute sequelae as outcomes |
| **Lazari et al (2023)** | Clinical markers of post-Chikungunya chronic inflammatory joint disease: A Brazilian cohort | Ineligible comparator group |
| **Carbonell-Estrany et al (2015)** | Long-Term Burden and Respiratory Effects of Respiratory Syncytial Virus Hospitalization in Preterm Infants—The SPRING Study | Self-reported outcomes (ineligible) |

### Appendix C. Supplementary Methods

#### Eligibility Criteria

The eligibility criteria were designed according to the Population, Intervention/Exposure, Comparator, Outcomes (PICO) framework. Eligible studies included patients with clinically diagnosed or laboratory-confirmed RSV, influenza, chikungunya, Zika, yellow fever, or dengue virus infection. Primary and recurrent infections were eligible, and studies reporting multi-organ post-infectious sequelae were included. Pre-clinical animal studies were excluded. Comparator groups included individuals without confirmed infection, including SARS-CoV-2-infected individuals where applicable. We included non-randomized cohort, case-control, and surveillance studies with appropriate comparator groups and excluded case reports, modelling studies, reviews, and other non-original articles. The study population included patients with clinically diagnosed and laboratory-confirmed infections with RSV, influenza, chikungunya virus, Zika virus, YFV, or dengue virus. Individuals with prior infection or re-infection were eligible for inclusion, and studies reporting multi-organ post-infectious sequelae (e.g., ICD-defined outcomes) were considered. We excluded pre-clinical studies in animal models. Eligible comparators included individuals without confirmed RSV, influenza, dengue, chikungunya, yellow fever, or Zika virus infection, with some studies additionally including SARS-CoV-2-infected individuals as an alternative comparison group. Comparator groups consisting of uninfected individuals but previously vaccinated against COVID-19 or influenza were included, as they represent a large proportion of the population and excluding them would not accurately reflect real-world dynamics. We included only non-randomized cohort, case-control, and surveillance study designs as long as they included adequate comparator groups, and we excluded case reports/case studies, systematic reviews, modelling studies, and non-original articles (e.g., reviews, commentaries, editorials, and opinion pieces). Studies conducted in diverse global healthcare settings (community, outpatient, and inpatient) across both high-income and low- and middle-income countries were eligible, and evidence from both pediatric and adult populations was included.

#### Search Strategy and Data Extraction

The search combined controlled vocabulary and free-text keywords related to the six viral pathogens (e.g., “Influenza, Human”, “RSV Infection”) with terms describing post-acute sequelae and epidemiological outcomes (e.g., “incidence”, “odds ratio”). No restrictions on date, language, or geographical setting were applied, and there was no need to contact authors. The full search strategies for all databases are provided in eAppendix A. Duplicates were removed prior to title/abstract and full-text screening. Full texts of potentially eligible articles were then retrieved and assessed against the predefined inclusion and exclusion criteria. Studies focusing on SARS-CoV-2/COVID-19 were excluded because PASC have already been extensively investigated, with numerous primary studies and systematic reviews published; therefore, this review intentionally focused on other viral pathogens for which evidence on post-acute sequelae is comparatively limited.

Most studies identified were excluded because they were unrelated to the six illnesses of interest, post-acute sequelae, or did not meet our study design criteria, as determined from titles or abstracts. Sixty-five studies underwent full-text review, with further exclusions due to ineligible study designs (Figure 1), absence of relevant outcomes (e.g., self-reported symptoms such as wheezing or fatigue), or inappropriate comparator groups (e.g., comparisons within the same exposure group by disease severity). Eight studies met the inclusion criteria from the primary search. We then conducted a forward citation search of these studies to identify additional relevant articles, repeating this process iteratively until no further eligible studies were found. For all included studies, we extracted data on study characteristics (authors, year, country, study period), population characteristics (e.g., age, health status), exposure and outcome definitions, study design, follow-up duration, sample size, cohort construction, and reported effect measures and estimates with corresponding confidence intervals.

The inclusion of six distinct viral pathogens and multiple outcomes made it difficult to capture all relevant studies within a single set of search terms per database. Therefore, we also performed an exhaustive forward citation search by screening the titles and abstracts within the reference lists of each included study. When additional eligible studies were identified through citation searching, we repeated this process for their reference lists until no further studies meeting our criteria were found. This yielded an additional 43 studies for full-text review and data extraction. For each of these additional studies, the same data were extracted as for the initial 8 studies found.

#### Addressing Appropriateness and Risk of Bias

Each of the six diseases were treated as the exposure, and diagnosis or symptoms of post-acute sequelae, all-cause hospitalization, or death as outcomes. The possible categories of risk of bias classification are low, some concerns, high, and very high risk of bias. Prior to the main risk of bias assessment, we completed assessments of appropriateness for each study, also within the ROBINS-E tool, to determine whether studies employed appropriate study designs, exposure measurements, study periods, follow-up time, and exposure-outcome models to report relevant effect measures. In the context of this systematic review, relevant effect measures include hazard ratios, odds ratios, incidence rate ratios, relative risks, and risk differences. The seven domains of bias primarily assess potential sources of bias arising from confounding, exposure measurement, participant selection, post-exposure interventions, missing data, outcome measurement, and selective reporting of results.

#### Meta-analysis

Given substantial heterogeneity across studies in study design and reporting, we attempted to standardize included estimates as much as possible prior to pooling. Sources of heterogeneity included differences in study populations (hospitalized inpatients, prolonged hospitalization, outpatients, or recorded cases), sample demographics (children, adults, older adults, or mixed populations), outcome definitions, such as diagnosis-based outcomes identified using ICD codes versus laboratory test-based outcomes, and definitions of the post-acute periods. When studies reported multiple post-acute follow-up windows, we selected estimates that most closely aligned with the post-acute definition used by the majority of studies, such as prioritizing follow-up beginning approximately 4 weeks or 30 days after exposure for influenza, where available. For studies that reported results for outpatients and patients with more severe hospitalization, we only included regular hospitalization effect estimates in meta-analyses, as this was the standard definition of severity common across all studies. Studies that included different populations (e.g., children or adults only) were pooled together due to the limited number of comparable effect estimates.

Some studies reported multiple effect estimates derived from distinct populations or subgroups without providing an overall pooled estimate. In such cases, we conducted separate meta-analyses for each eligible population-specific estimate but included no more than one estimate per dataset within any single meta-analysis to avoid unit-of-analysis errors, which would artificially reduce the observed variance and bias the pooled estimate.

Meta-analyses were performed only when at least three independent studies reported comparable exposure-outcome models using the same effect measure. When fewer than three studies were available for a given exposure-outcome and effect measure combination, results were summarized descriptively rather than pooled.

**Table S1:** Inclusion and exclusion criteria for study selection.

| Inclusion Criteria | Exclusion Criteria |
| --- | --- |
| • Studies on adult, pediatric and geriatric populations<br>• Patients with clinically-confirmed infections or via laboratory testing<br>• Patients previously infected with influenza, RSV, dengue, Yellow Fever, Chikungunya, Zika viruses<br>• Patients re-infected with influenza, RSV, dengue, yellow fever, chikungunya, Zika viruses<br>• Patients with post-acute multi-organ sequelae as recognized under the International Classification of Diseases<br>• Post-acute sequelae that occur > 4 weeks after the acute infection<br>• Cohort, case-control, surveillance studies | • Patients vaccinated against the aforementioned infections<br>• Patients with pre-existing history of the condition (the post-acute sequelae) being studied<br>• Studies describing self-reported symptoms of post-acute sequelae<br>• Studies describing the effectiveness of vaccines against the aforementioned viral infections<br>• Animal subjects<br>• Case studies, systematic reviews, case reports, letters, book chapters, conference abstracts, commentaries, editorials |

**Figure S1:**
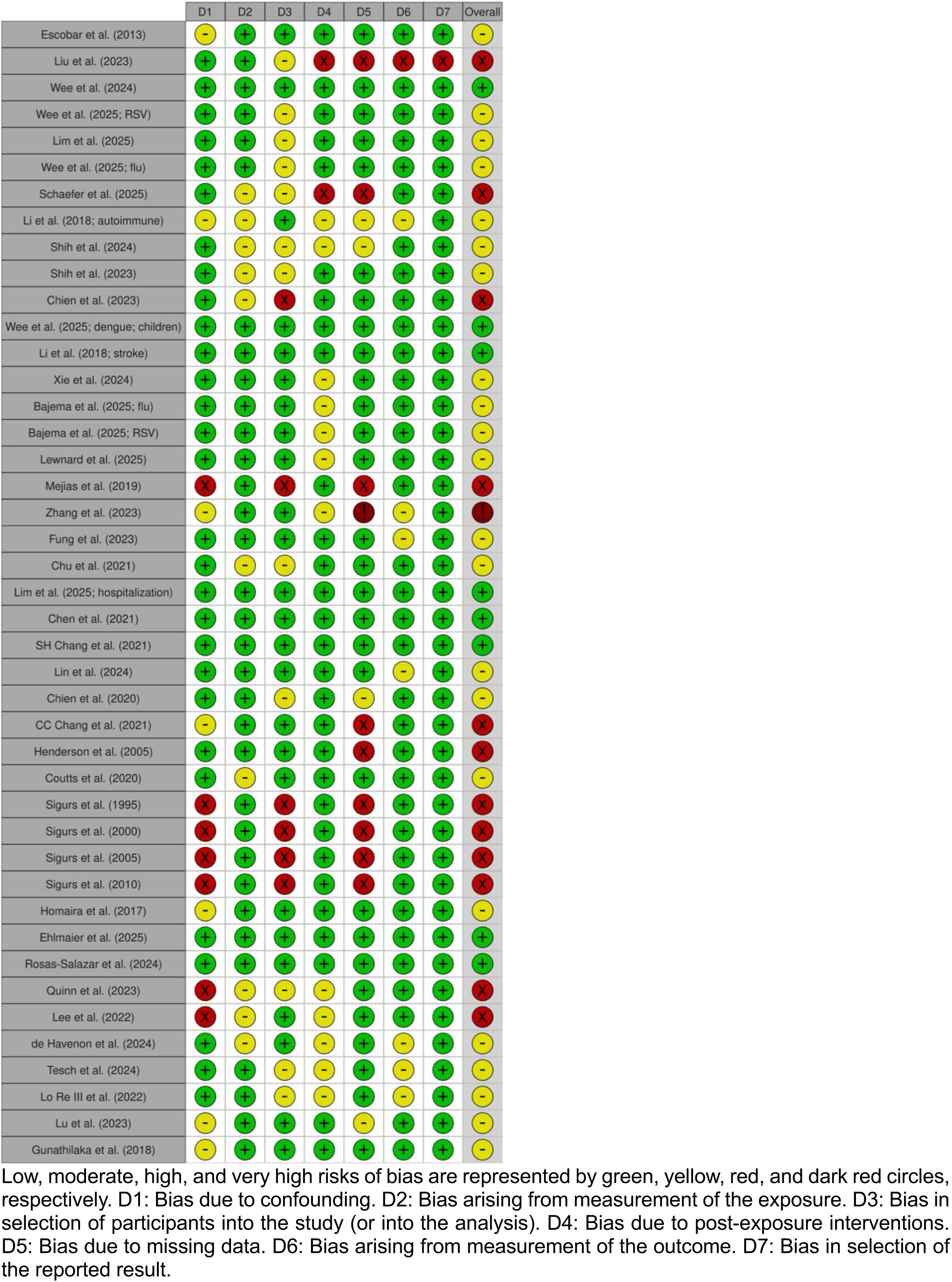
Traffic light chart of individual study risks of bias per domain and overall.

**Figure S2:**
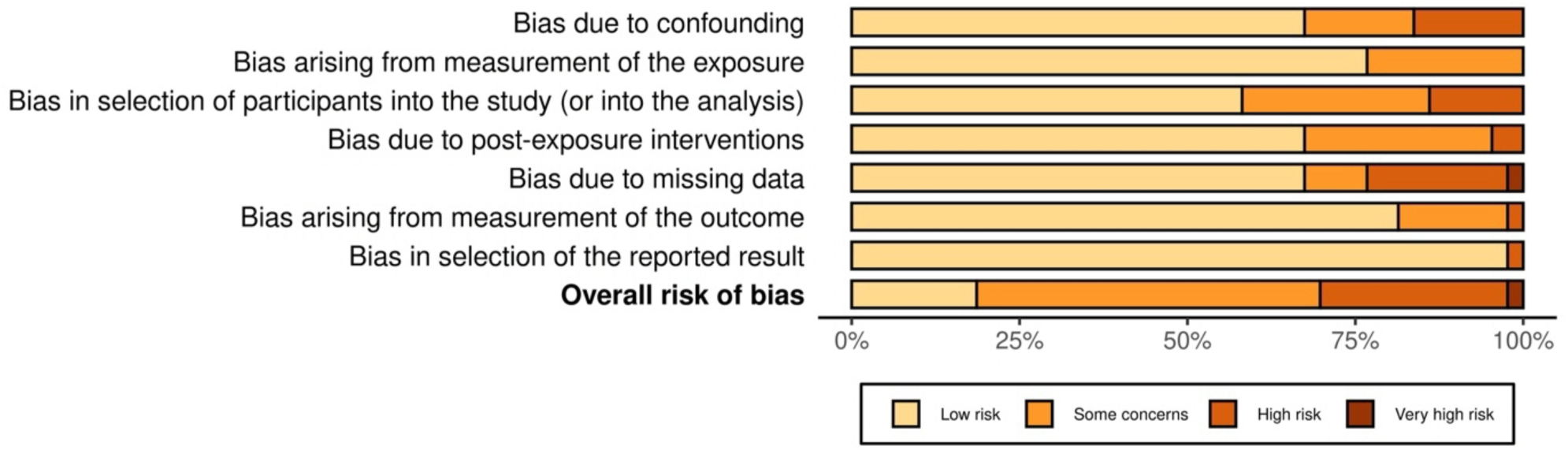
Bar charts of the distribution of risks of bias across the 7 domains.

**Figure S3:**
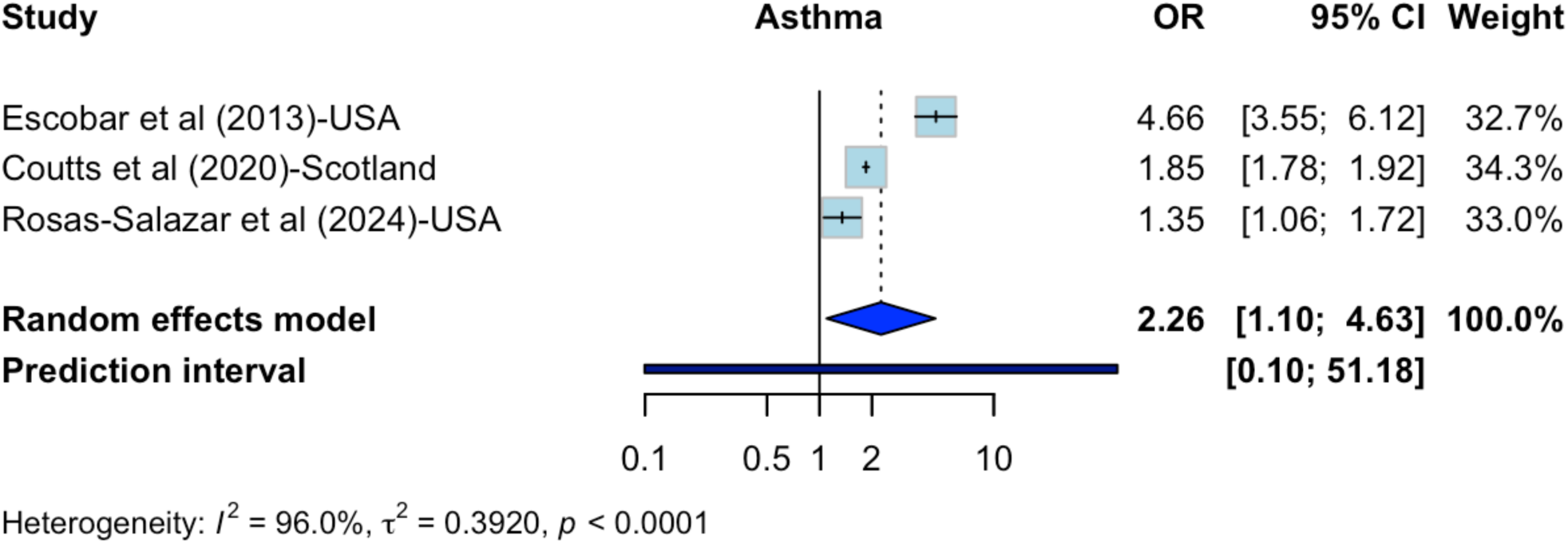
Sensitivity meta-analysis for RSV-asthma.

**Figure S4:**
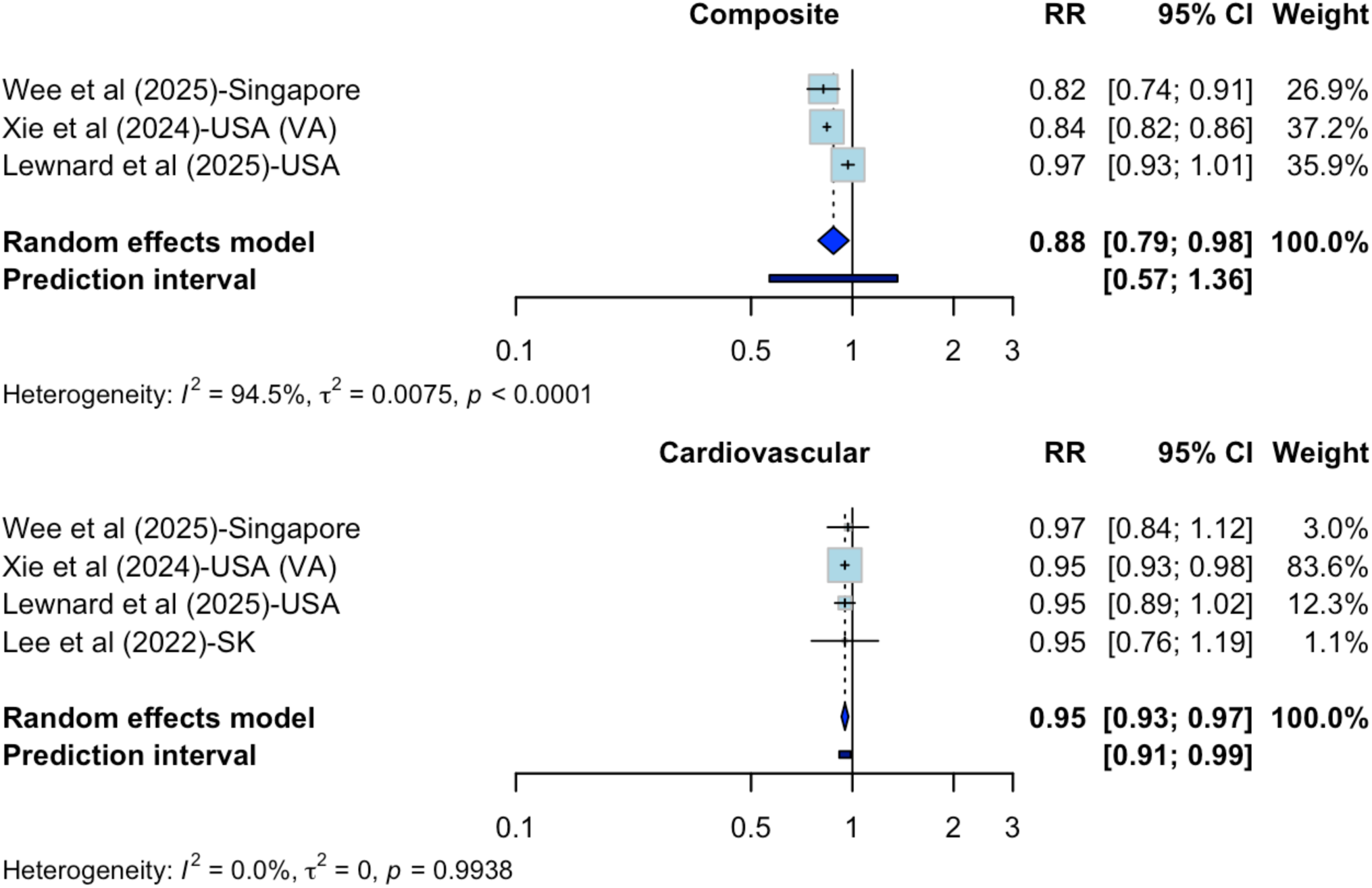
Sensitivity meta-analyses for influenza and any post-acute sequelae and cardiovascular outcomes.

**Figure S5:**
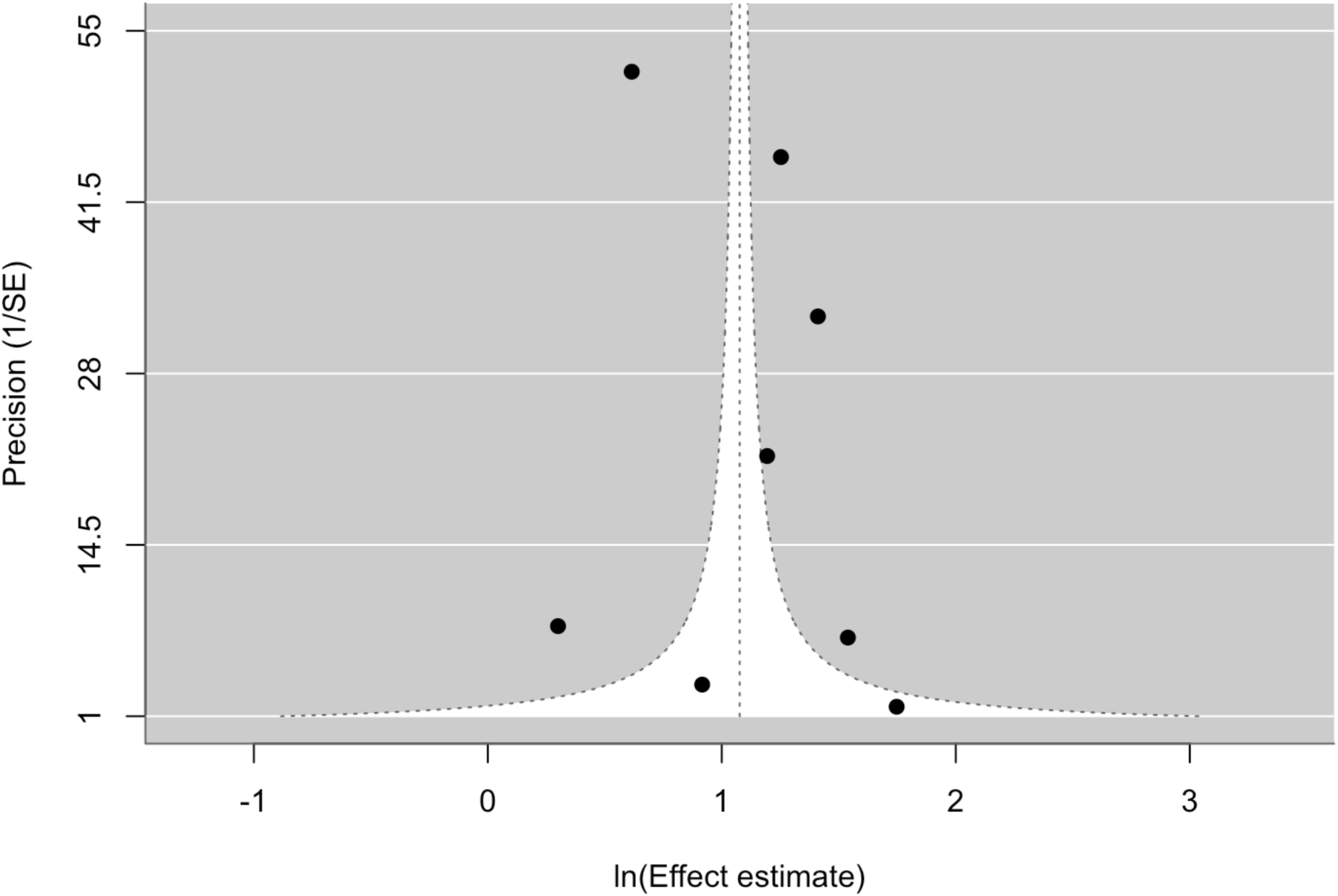
Publication bias funnel plots for RSV-asthma.

**Figure S6:**
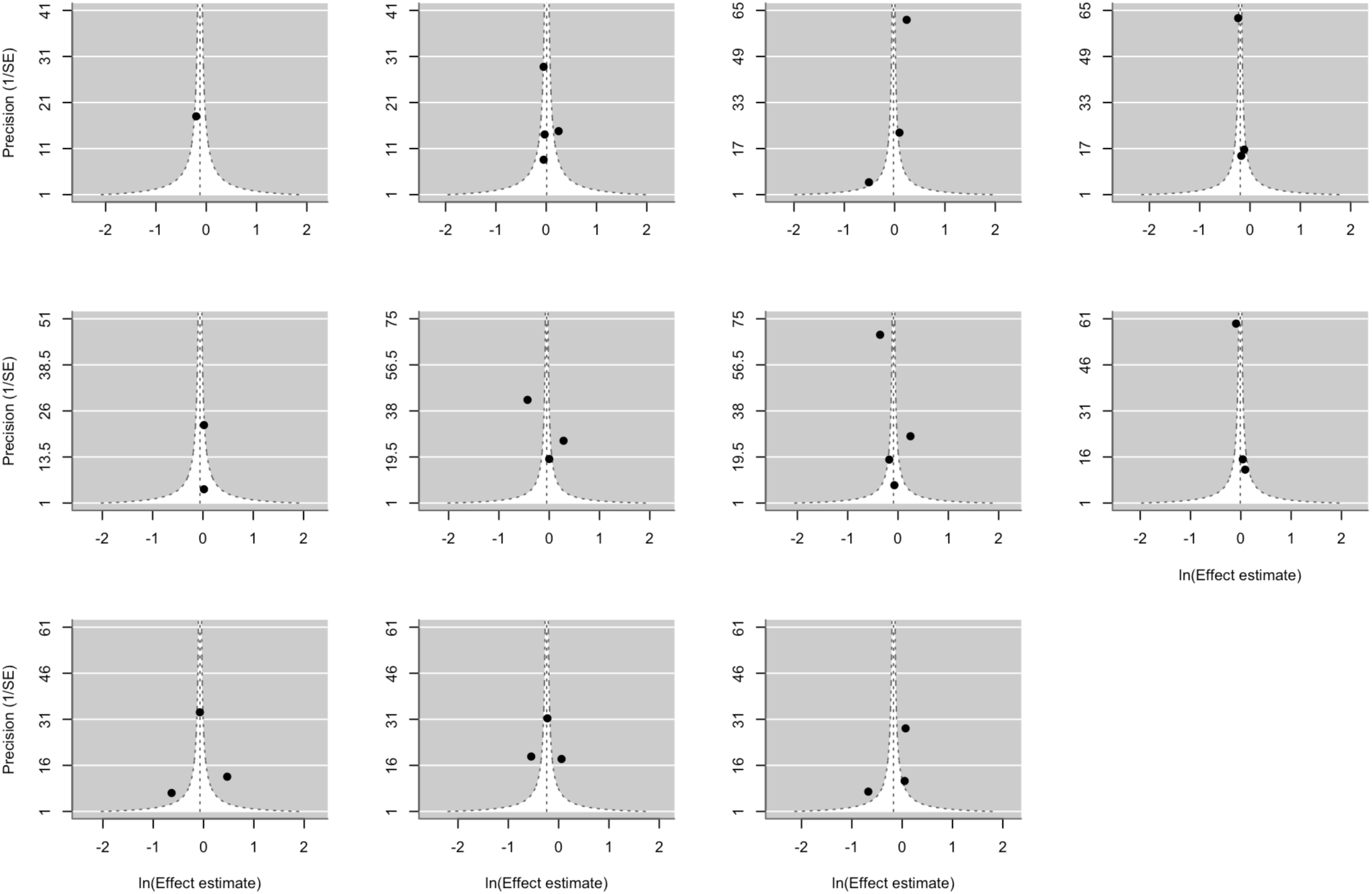
Publication bias funnel plots for influenza and its meta-analyzed outcomes.

**Figure S7:**
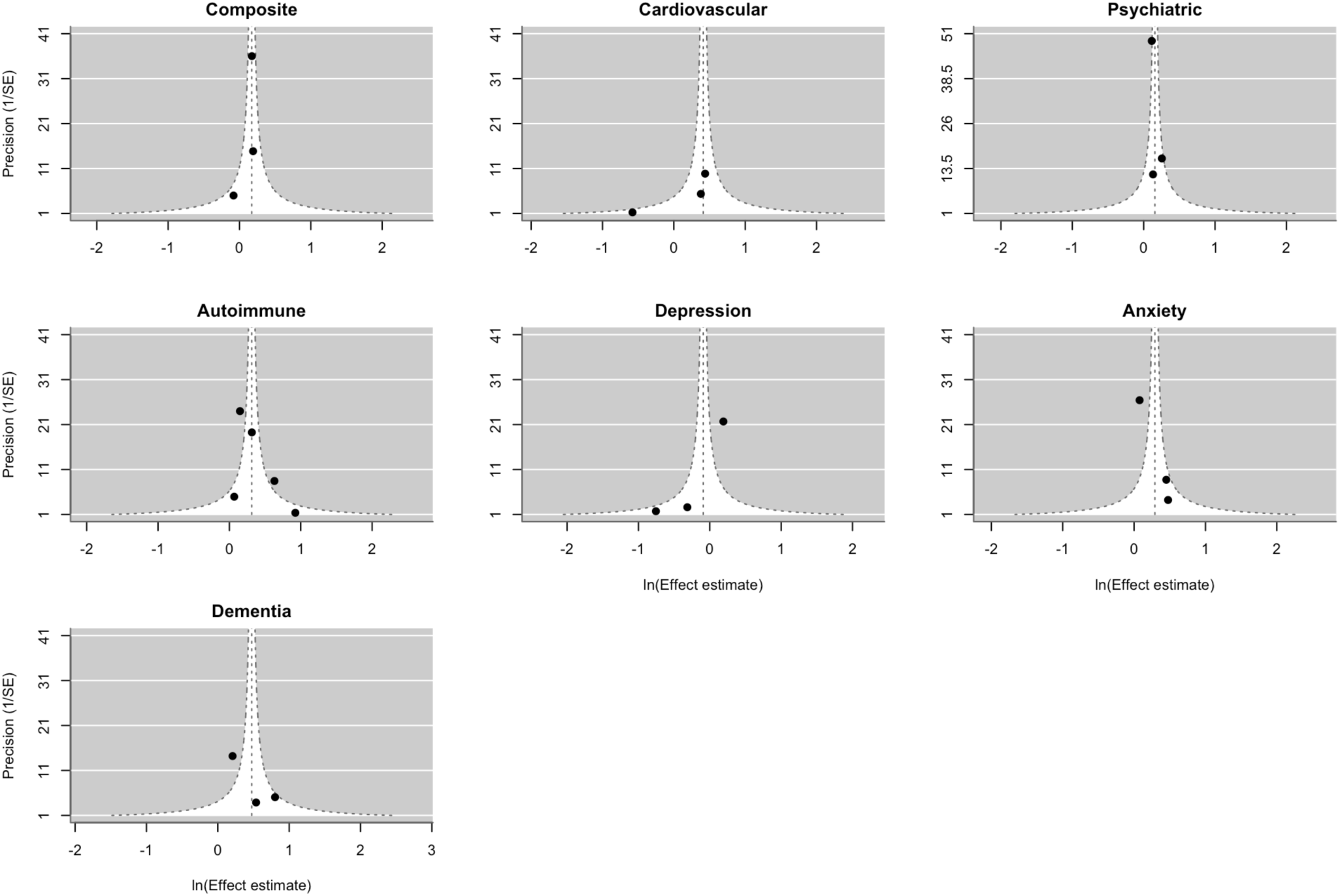
Publication bias funnel plots for dengue and its meta-analyzed outcomes.

